# Adverse Cardiovascular Complications Following Prescription of Programmed Cell Death 1 (PD-1) and Programmed Cell Death Ligand 1 (PD-L1) Inhibitors: A Propensity-Score Matched Cohort Study with Competing Risk Analysis

**DOI:** 10.1101/2020.12.21.20248648

**Authors:** Jiandong Zhou, Sharen Lee, Ishan Lakhani, Lei Yang, Tong Liu, Yuhui Zhang, Yunlong Xia, Wing Tak Wong, Kelvin King Hei Bao, Ian Chi Kei Wong, Gary Tse, Qingpeng Zhang

## Abstract

**Background:** Programmed death-1 (PD-1) and programmed death-ligand 1 (PD-L1) inhibitors, such as pembrolizumab, nivolumab and atezolizumab, are major classes of immune checkpoint inhibitors that are increasingly used for cancer treatment. However, their use is associated with adverse cardiovascular events. We examined the incidence of new-onset cardiac complications in patients receiving PD-1 or PD-L1 inhibitors.

**Methods:** Patients receiving PD-1 or PD-L1 inhibitors since their launch up to 31^st^ December 2019 at publicly funded hospitals of Hong Kong, China, without pre-existing cardiac complications were included. The primary outcome was a composite of incident heart failure, acute myocardial infarction, atrial fibrillation or atrial flutter with the last follow-up date of 31^st^ December 2020. Propensity score matching between PD-L1 inhibitor use and PD-1 inhibitor use with a 1:2 ratio for patient demographics, past comorbidities and non-PD-1/PD-L1 medications was performed.

**Results:** A total of 1959 patients were included. Over a median follow-up of 247 days (interquartile range [IQR]: 72-506), 320 (incidence rate [IR]: 16.31%) patients met the primary outcome after PD-1/PD-L1 treatment: 244 (IR: 12.57%) with heart failure, 38 (IR: 1.93%) with acute myocardial infarction, 54 (IR: 2.75%) with atrial fibrillation, 6 (IR: 0.31%) with atrial flutter. Compared with PD-1 inhibitor treatment, PD-L1 inhibitor treatment was significantly associated with lower risks of the composite outcome both before (hazard ratio [HR]: 0.32, 95% CI: [0.18-0.59], P value=0.0002) and after matching (HR: 0.34, 95% CI: [0.18-0.65], P value=0.001), and lower all-cause mortality risks before matching (HR: 0.77, 95% CI: [0.64-0.93], P value=0.0078) and after matching (HR: 0.80, 95% CI: [0.65-1.00], P value=0.0463). Patients who developed cardiac complications had shorter average readmission intervals and a higher number of hospitalizations after treatment with PD-1/PD-L1 inhibitors in both the unmatched and matched cohorts (P value<0.0001). Competing risk analysis with cause-specific and subdistribution hazard models and multiple approaches based on the propensity score all confirmed these observations.

**Conclusions:** Compared with PD-1 treatment, PD-L1 treatment was significantly associated with lower risk of new onset cardiac complications and all-cause mortality both before and after propensity score matching.

## Introduction

The programmed death-1 (PD-1)/programmed death-ligand 1 (PD-L1) pathway is one of the major immune checkpoints for mitigating the immune response to prevent autoimmunity. However, cancer cells often devise strategies to hijack these mechanisms to evade anti-tumor immunity. In this regard, inhibitors of PD-1 (e.g., pembrolizumab, nivolumab, cemiplimab) and PD-L1 (e.g. atezolizumab, avelumab, durvalumab) have shown clinical efficacies against different types of solid tumors, including melanoma, non-small cell lung cancer (NSCLC), urothelial carcinoma and bladder cancer. Pembrolizumab is also the first agent to receive a “pan-cancer” approval by the United States Food and Drug Administration (FDA) for the treatment of unresectable or metastatic solid tumors that have high microsatellite instability or mismatch repair deficiency.

Despite their treatment efficacy in clinical oncology, immune-related adverse events associated with the use of immune checkpoint inhibitors (ICIs) are now increasingly recognized (1–4). Adverse events include atherosclerosis, colitis, hepatitis, adrenocorticotropic hormone insufficiency, hypothyroidism, type 1 diabetes mellitus, and acute kidney injury (5–7). To this end, cardiovascular complications are estimated to constitute approximately 2% of ICI-related adverse drug reactions (8). The commonest is myocarditis, but other cardiovascular abnormalities reported are left ventricular dysfunction, acute myocardial infarction (AMI), cardiac arrhythmias and heart failure (9). These cardiovascular complications typically present with clinical heterogeneity, and in turn account for the high morbidity and mortality rates observed in such patient cohorts. Whilst cardiotoxicity is being documented with an increasing frequency, their cumulative incidence rates remain largely unexplored. In this territory-wide study, we examined the incidence of cardiovascular events of incident heart failure, acute myocardial infarction, atrial fibrillation or atrial flutter in cancer patients receiving PD-1 or PD-L1 inhibitors.

## Methods

### Study Population

This study was approved by The Joint Chinese University of Hong Kong - New Territories East Cluster Clinical Research Ethics Committee. Patients receiving PD-1 or PD-L1 inhibitors since their launch up to 31^st^ December 2020 at publicly funded hospitals or their associated outpatient/ambulatory care facilities, without pre-existing cardiac complications were included. Patient data were obtained using the electronic health record database, which is connected to the territory-wide Clinical Data Analysis and Reporting System (CDARS). The system is an integrative centralized platform that permits the extraction of clinical data for analysis and reporting. The system attributes each patient a unique reference identification number, allowing for the retrieval of comprehensive medical records, including disease diagnoses, clinical comorbidities, laboratory parameters and operative procedures. Patients or the public were not involved in any aspect of this study. The system has been previously used by both our team and other teams in Hong Kong (10–12).

### Patient Data

The following clinical data were extracted: patient characteristics, including demographic details (baseline age and gender), specific pre-existing comorbidities before drug prescriptions, laboratory examinations (including complete blood counts, biochemical tests, lipid and glycemic profiles) were extracted. Past comorbidities from January 1^st^, 2013 to December 31^st^, 2020 were extracted, and categorized into hypertension, liver diseases, hip fractures/accident falls, renal diseases, diabetes mellitus, maligt dysrhythmia, chronic obstructive pulmonary disease, ischemic heart disease, peripheral vascular disease, endocrine diseases, gastrointestinal diseases, and stroke/transient ischemic attack. The International Classification of Disease, Ninth Edition (ICD-9) codes that were used to extract the specific comorbidities and outcomes are included in **Supplementary Table 2**. The overall dosage and accumulative duration for patients with new-onset cardiac complications are reported.

### Primary outcomes on follow-up

The primary outcome was a composite of incident heart failure, acute myocardial infarction, atrial fibrillation, and atrial flutter. The follow-up period was defined as the first PD-1/PD-L1 prescription date until the primary endpoint or death occurred, or until the end date of August 31^st^, 2020, whichever was earlier.

### Statistical analysis

Continuous variables were presented as median (95% confidence interval [CI] or interquartile range [IQR]) and categorical variables were presented as count (%). The Mann-Whitney U test was used to compare continuous variables. The χ^2^ test with Yates’ correction was used for 2×2 contingency data, and Pearson’s χ^2^ test was used for contingency data for variables with more than two categories. The patients with PD-L1 were matched with PD1 controls through propensity score matching of 1:2 ratio, based on patient demographics, Charlson standard comorbidity index, past comorbidities, and non-PD-1/PD-L1 medications. Negligible post-weighting intergroup standardized mean difference (SMD) was defined as SMD < 0.2. To identify the important predictors associated with new-onset cardiac complications of patients after PD-1/PD-L1 treatment, univariate Cox regression was used to calculate hazard ratios (HRs) and 95% CIs. In addition to propensity score matching, the following approaches based on the propensity scores were employed: propensity score stratification (13), inverse probability weighting (14), and high-dimensional propensity score adjustment (15). Paired hospitalization characteristics of patients before and after treatment were compared both in the unmatched and matched cohorts. A two-sided α of < 0.05 was considered statistically significant. Statistical analyses were performed using RStudio software (Version: 1.1.456) and Python (Version: 3.6).

## Results

### Baseline characteristics

Initially, 2426 cancer patients receiving PD-1/PD-L1 inhibitors were identified (Figure 1). In total 1959 patients remained in the study cohort after excluding 433 patients with prior cardiac complications and 34 patients who received both PD1 and PDL1 treatments. Propensity score matching with 1:2 ratio between PD-1 and PD-L1 inhibitor use based on demographics, Charlson standard comorbidity index, prior comorbidities, and non-PD-1/PD-L1 medications was performed. This yielded a matched cohort of 663 patients (Table 1).

**Figure 1.**
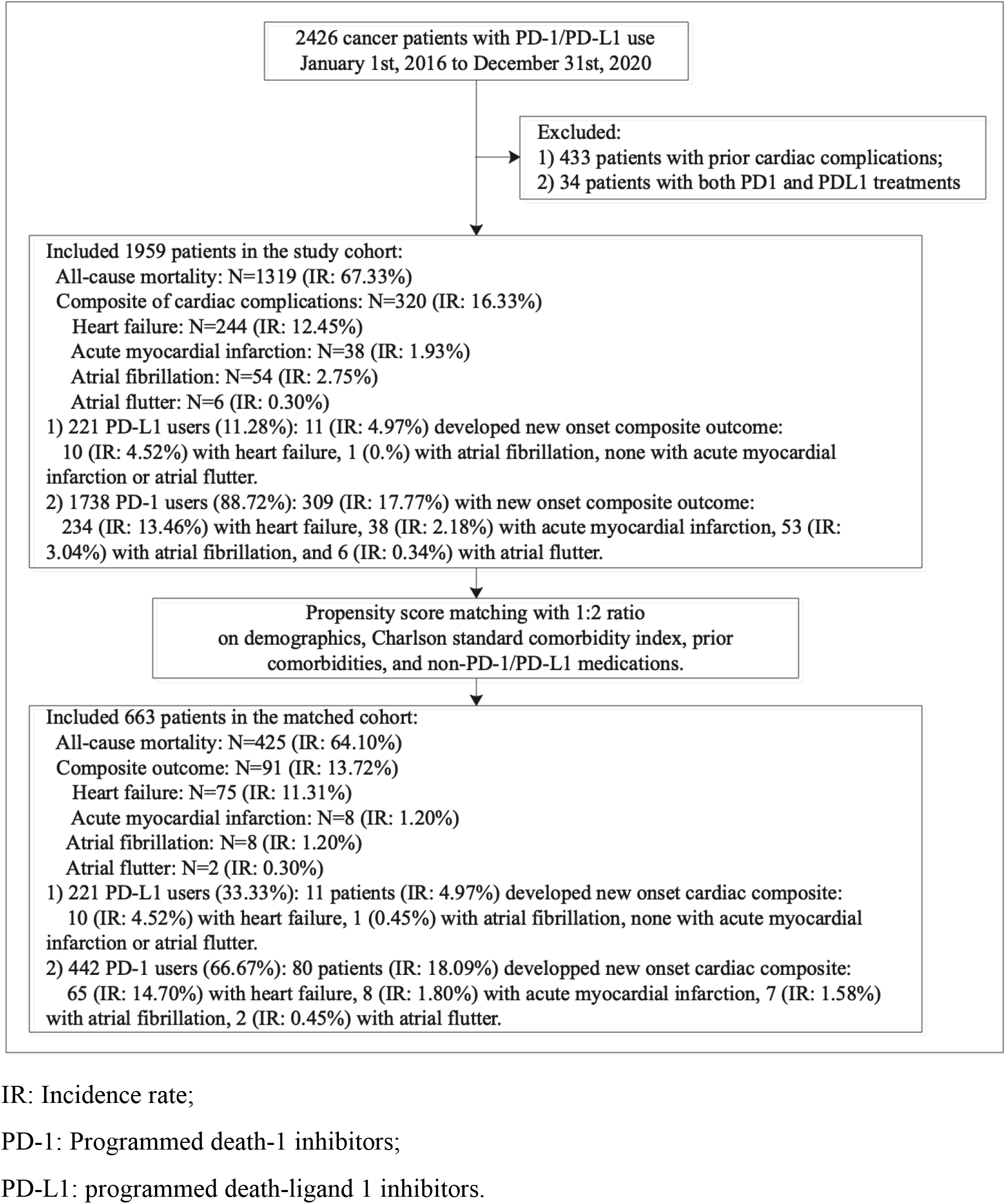
Study flow diagram describing derivation of the study cohort.

**Table 1.**
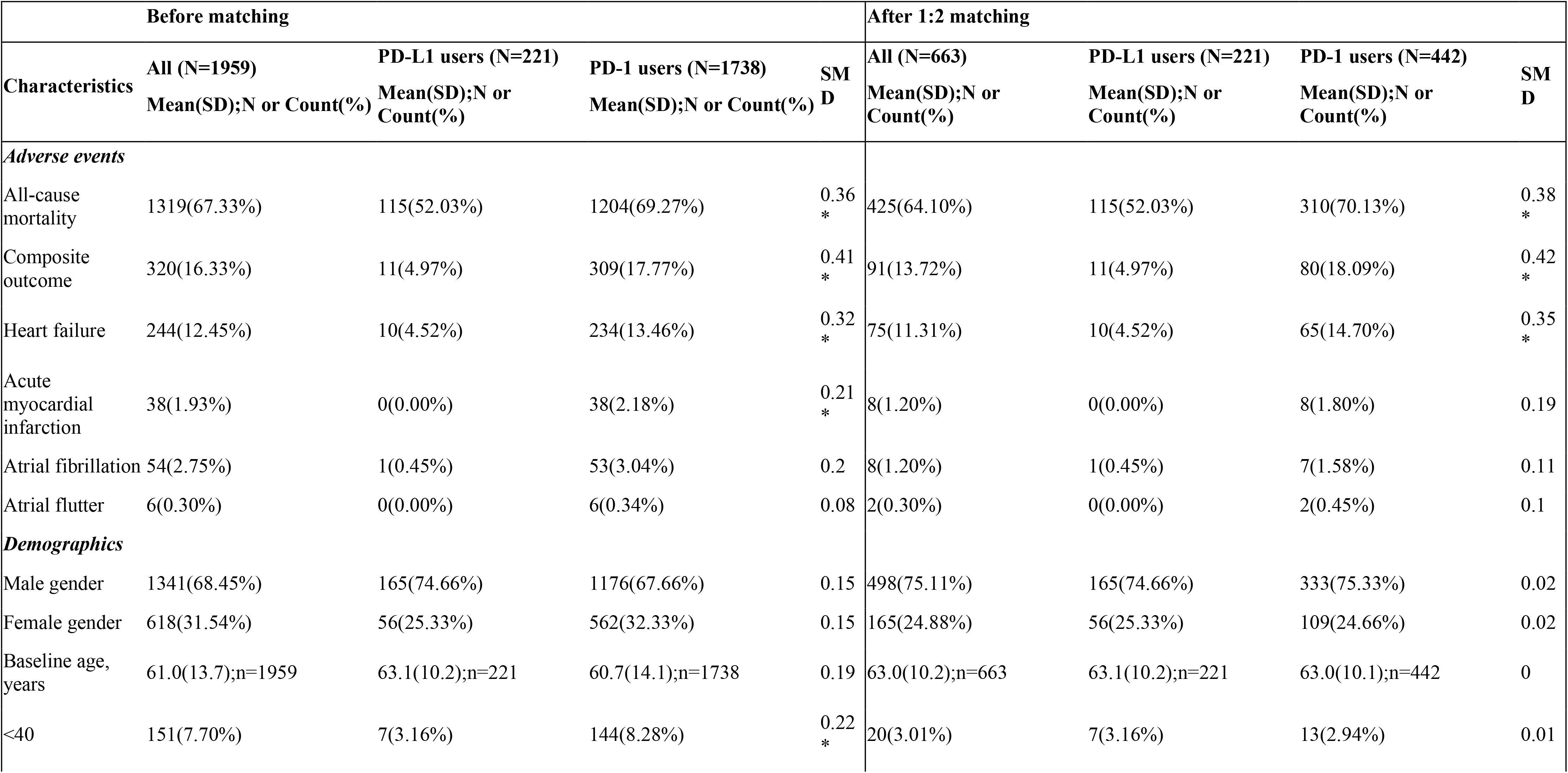
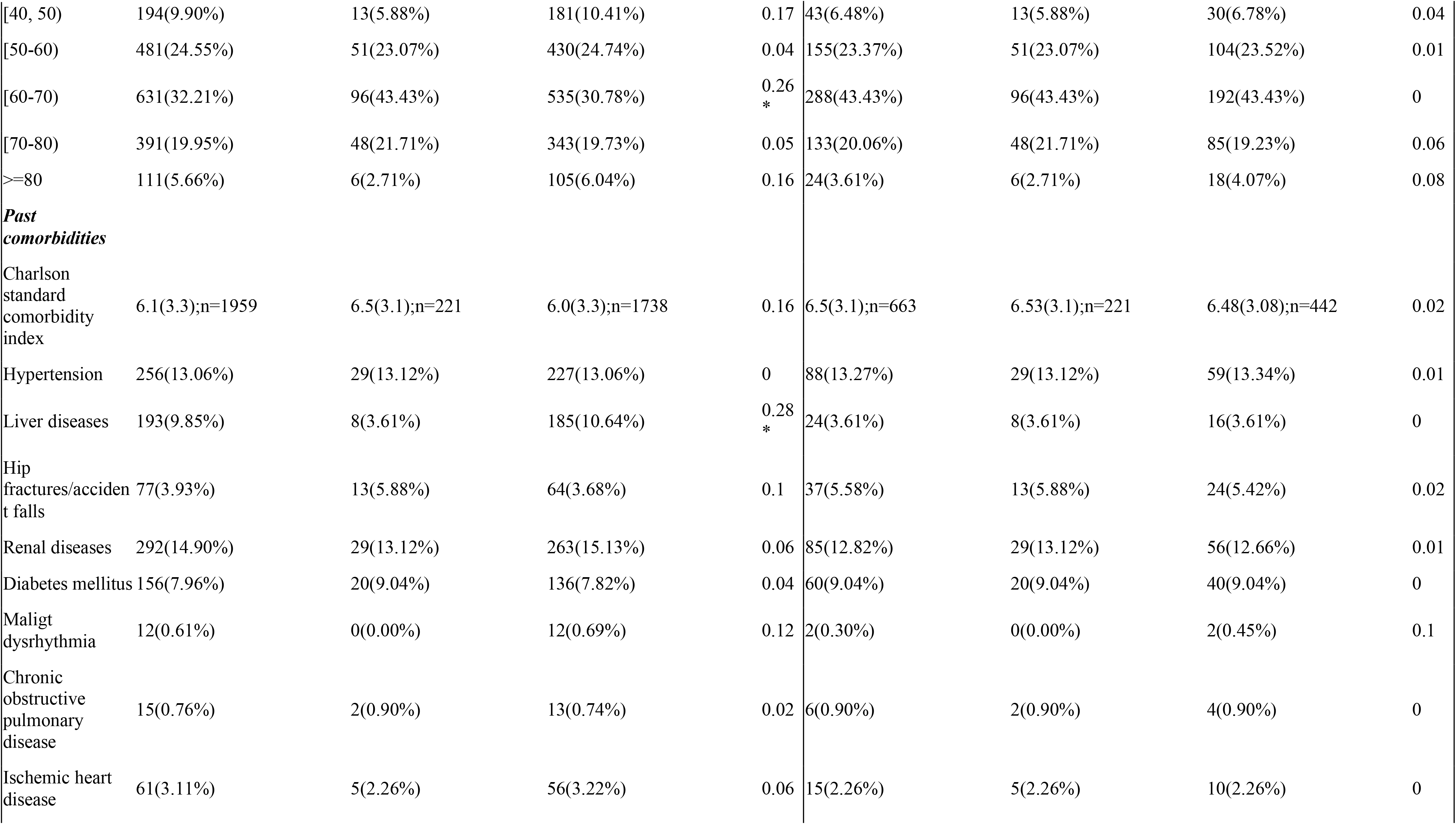
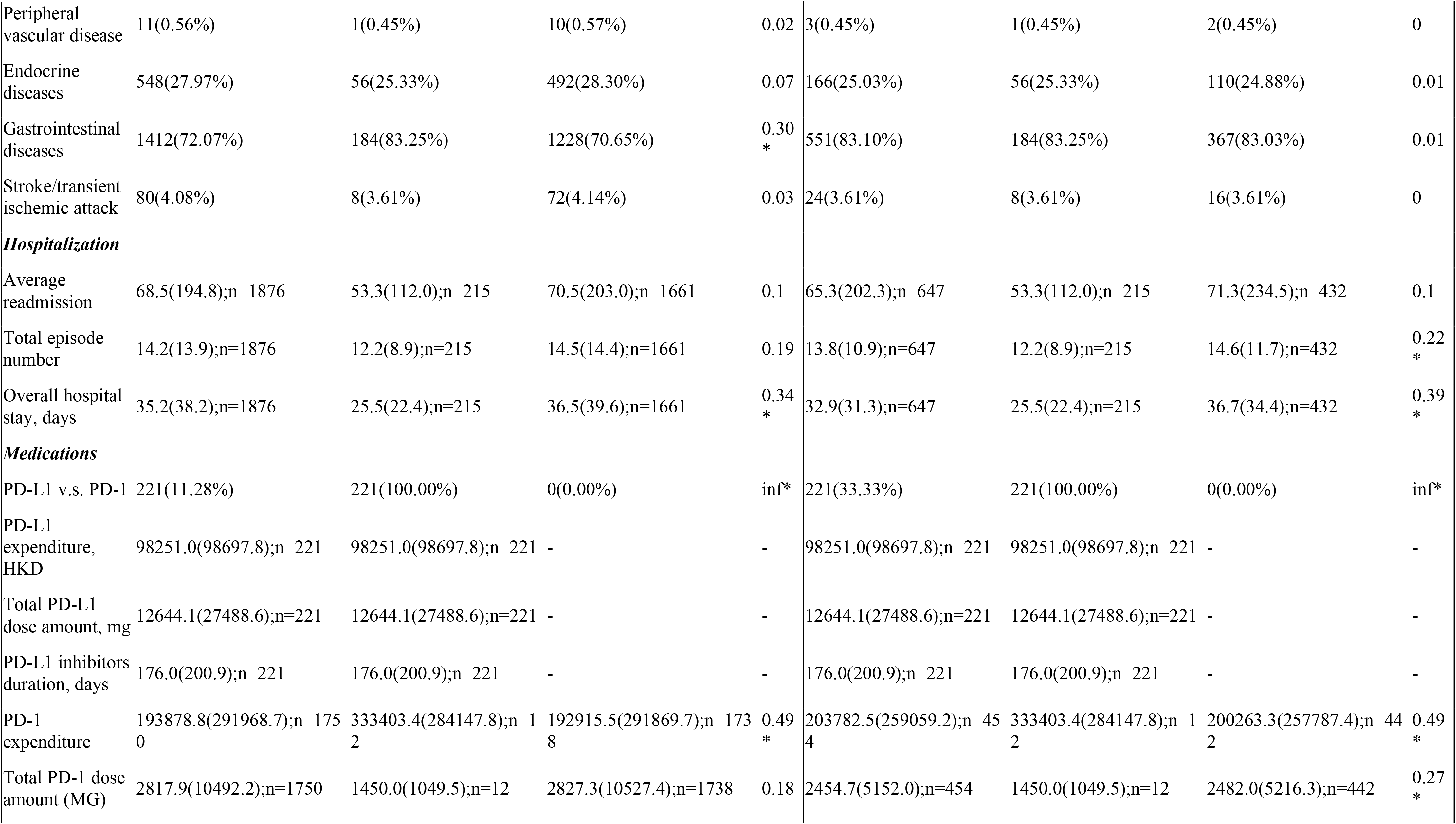
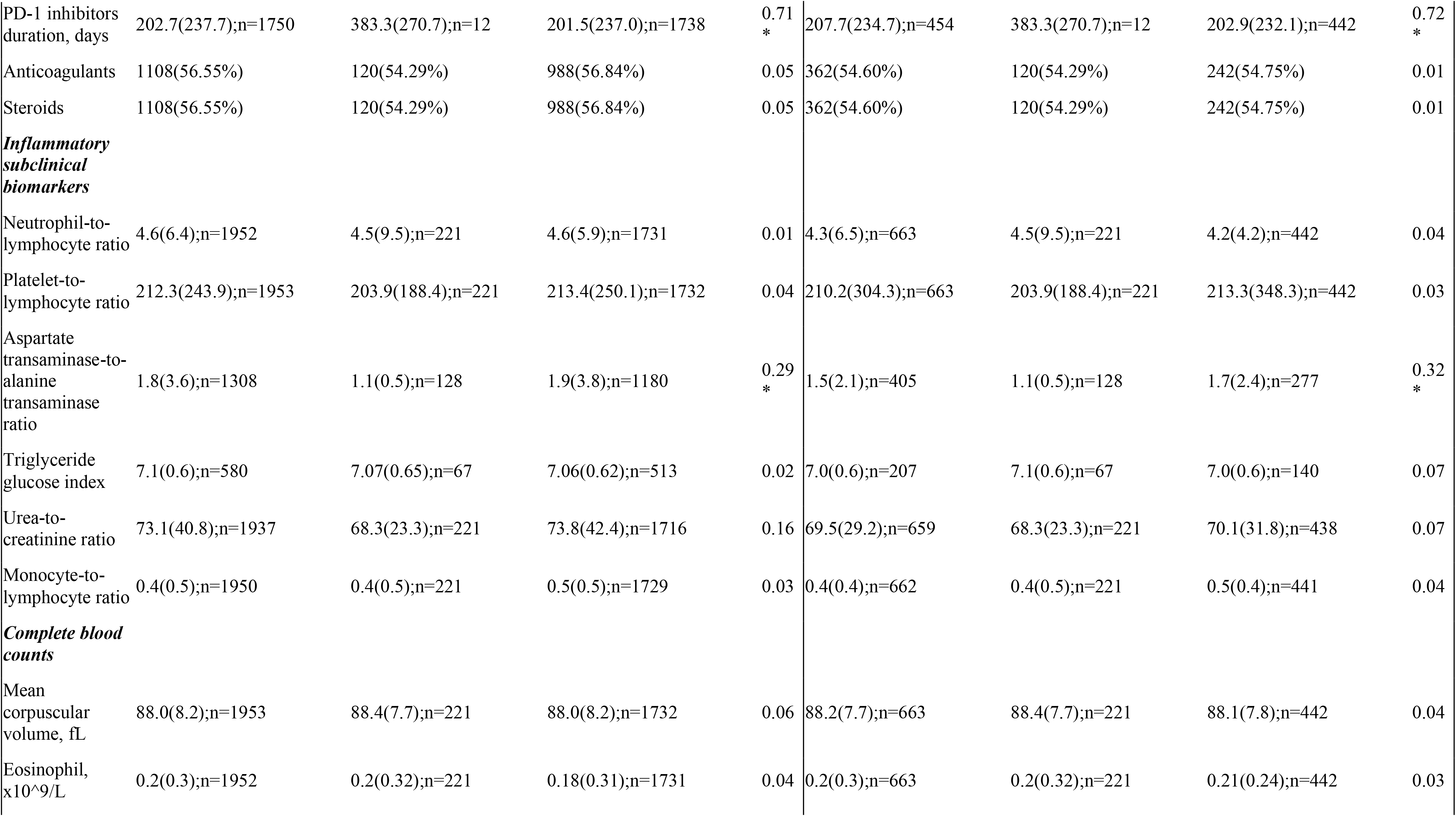
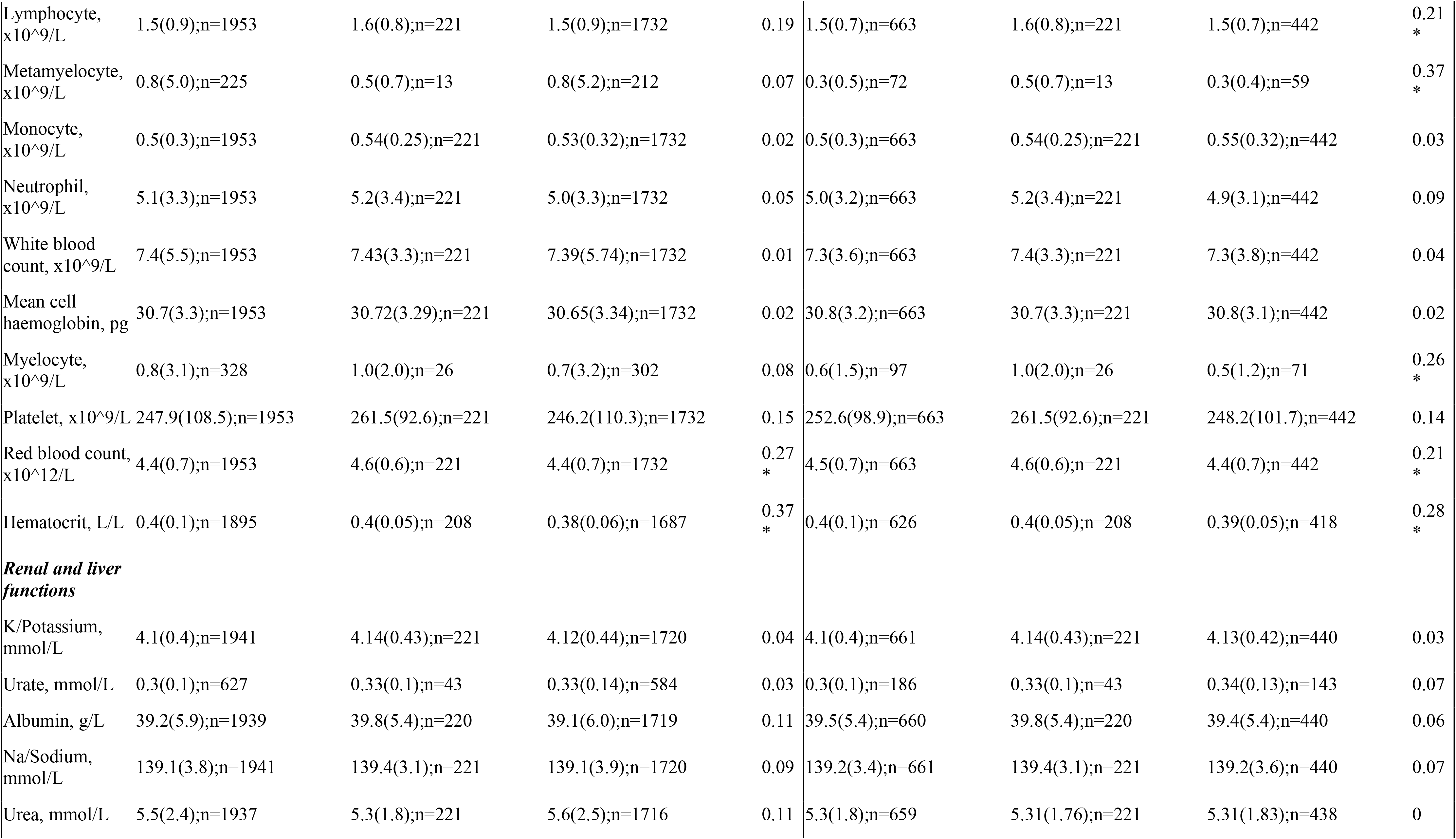
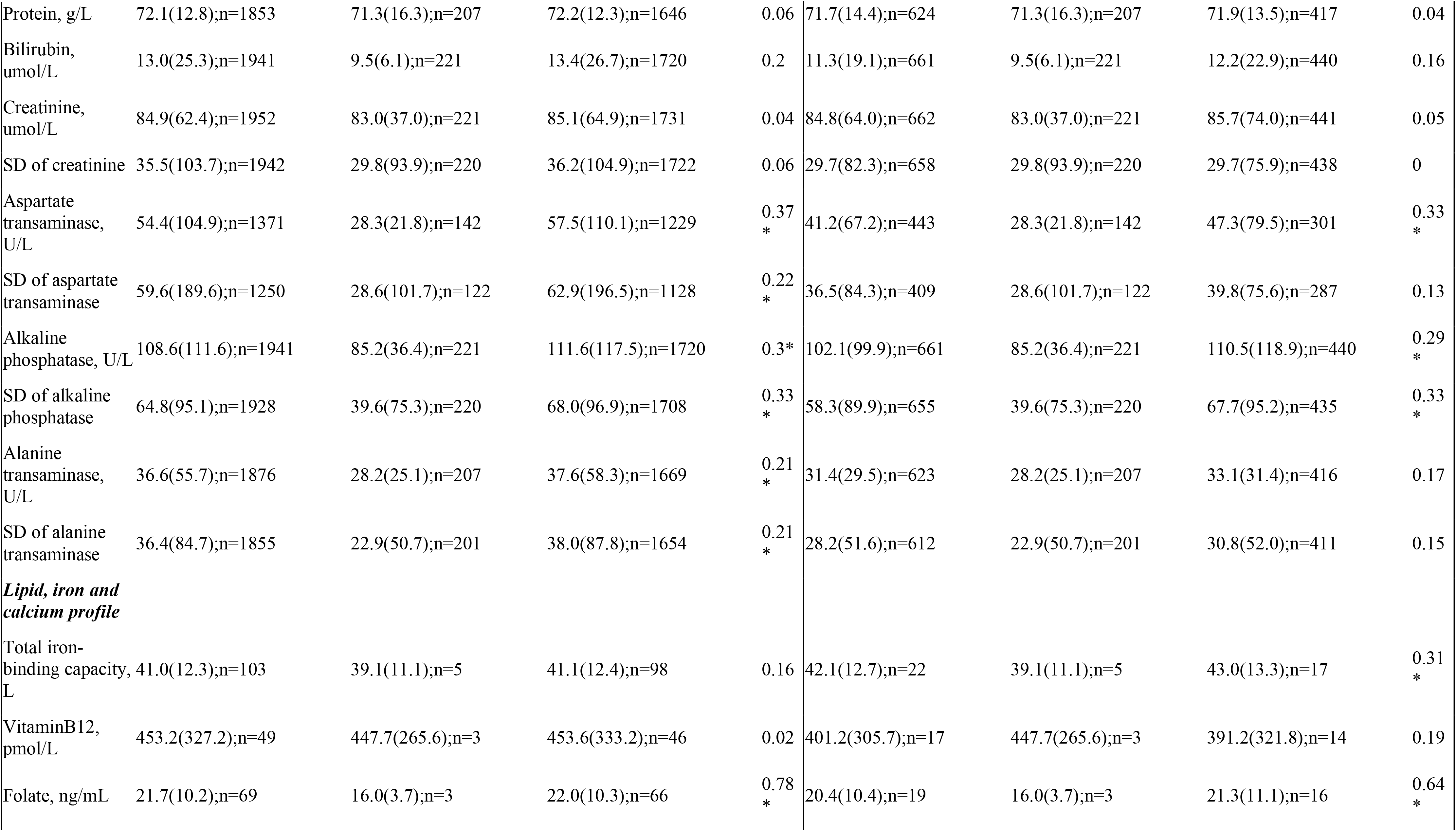
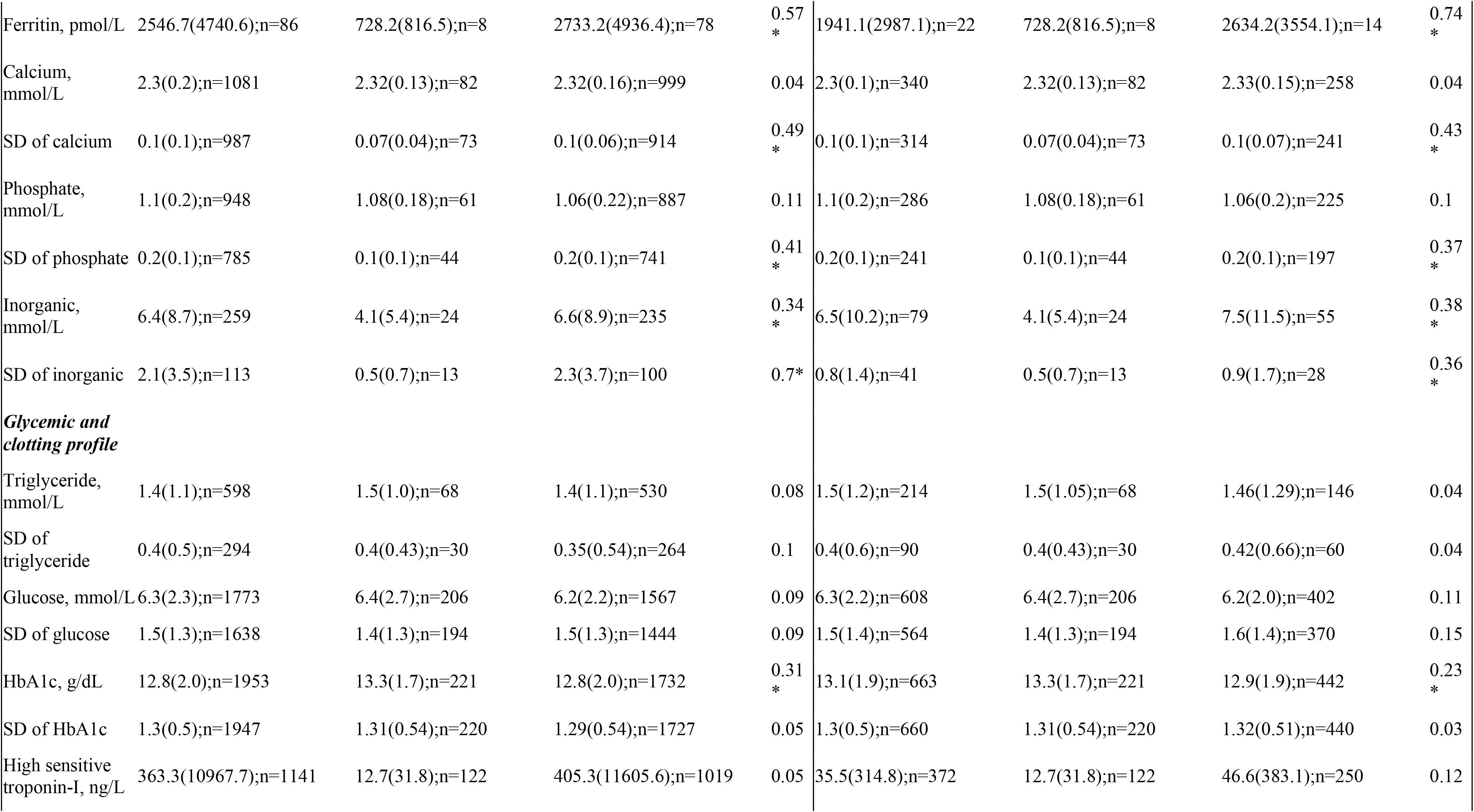
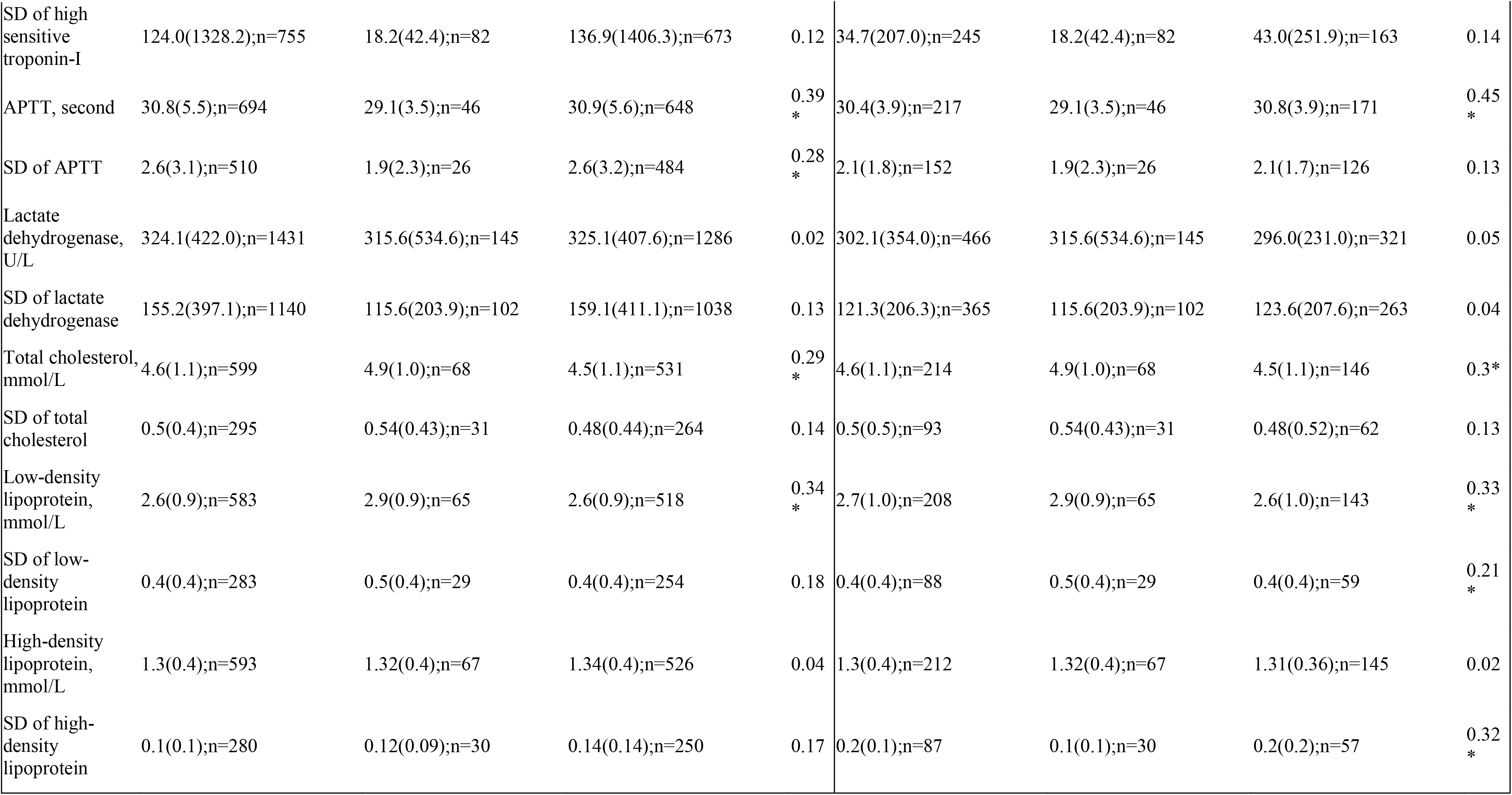
Clinical characteristics of patients with PD-1 use and PD-L1 use before and after 1:2 propensity score matching. * for *SMD* 0.2 APTT: applied partial thromboplastin test; PD-1: Programmed death 1 inhibitors; PD-L1: programmed death 1 ligand inhibitors

Propensity score matching in 1:2 ratio between PD-1 users and PD-L1 users using the nearest neighbor search strategy was used. The results of logistics regression for potential confounders used in propensity score calculations, balance between groups, and estimations of bootstrapped standard error are shown in **Supplementary Table 3, 4 and 5**, respectively. Distributions of propensity scores before and after matching are shown in **Supplementary Figure 1**. These results indicate that the covariates between the groups are balanced after matching.

### Adverse cardiovascular outcomes on follow-up and their significant predictors

In the matched cohort, 425 (IR: 64.10%) patients died and 91 (IR: 13.72%) developed new onset cardiac composite outcome. Amongst the latter, 75 (IR: 11.31%) developed heart failure, 8 (IR: 1.20%) developed acute myocardial infarction, 8 (IR: 1.20%) developed atrial fibrillation and 2 (IR: 0.30%) developed atrial flutter. The incidence rate of the composite outcome was lower in the PD-L1 cohort than in the PD-1 cohort (7.0% vs. 20.7%; P<0.001).

In the 221 PD-L1 users, there were 11 patients (IR: 4.97%) who developed the composite outcome, in which 10 (IR: 4.52%) with heart failure, 1 (0.45%) with atrial fibrillation, but none with acute myocardial infarction or atrial flutter. In the 442 patients PD-1 users, 80 patients (IR: 18.09%) developed the composite outcome. Of the latter group, 65 (IR: 14.70%) developed heart failure, 8 (IR: 1.80%) developed acute myocardial infarction, 7 (IR: 1.58%) developed atrial fibrillation, and 2 (IR: 0.45%) developed atrial flutter.

The breakdown on the individual adverse events is shown in Figure 1 and the patient characteristics stratified by adverse cardiovascular outcomes are shown in Table 2 and Table 3. The Kaplan-Meier survival curves and the cumulative incidence curves of new onset cardiac complications and all-cause mortality in cancer patients stratified by PD-1 or PD-L1 inhibitor use before and after 1:2 propensity score matching were presented in Figure 2 and Figure 3, respectively. The baseline characteristics of the cohort stratified by mortality status are shown in **Supplementary Table 6**.

**Figure 2.**
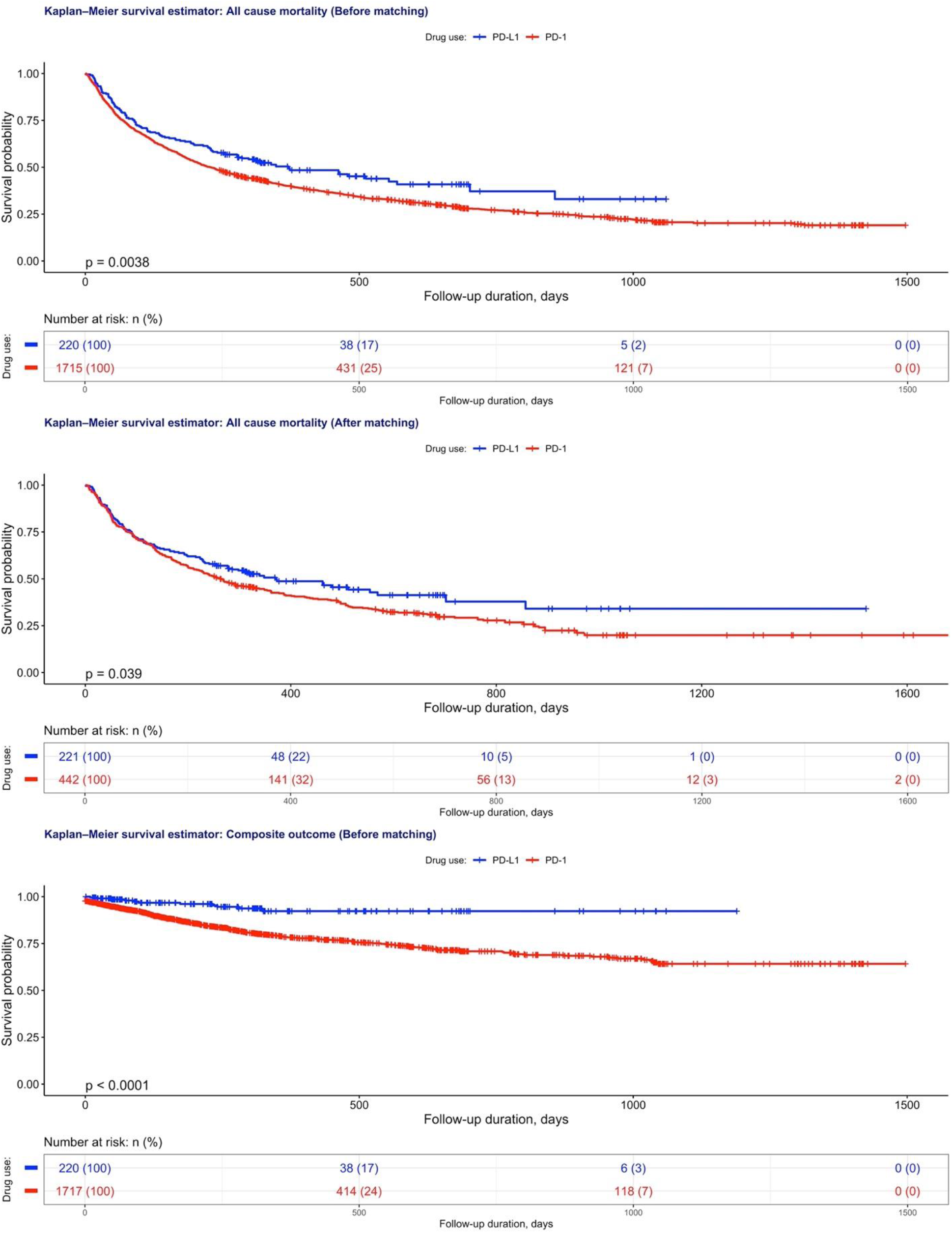
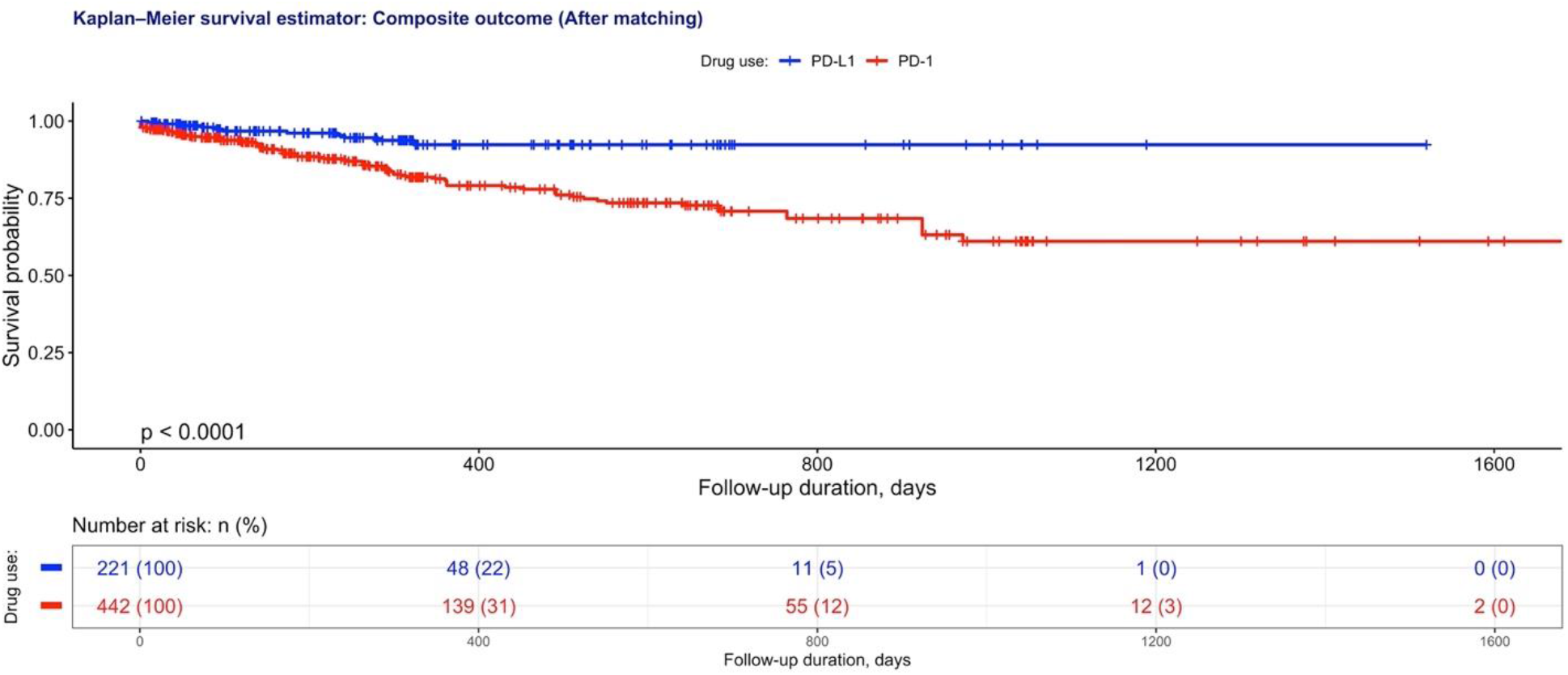
Kaplan-Meier survival curves of new onset cardiac complications and all-cause mortality in cancer patients stratified by PD-1 or PD-L1 inhibitor use before and after 1:2 propensity score matching.

**Figure 3.**
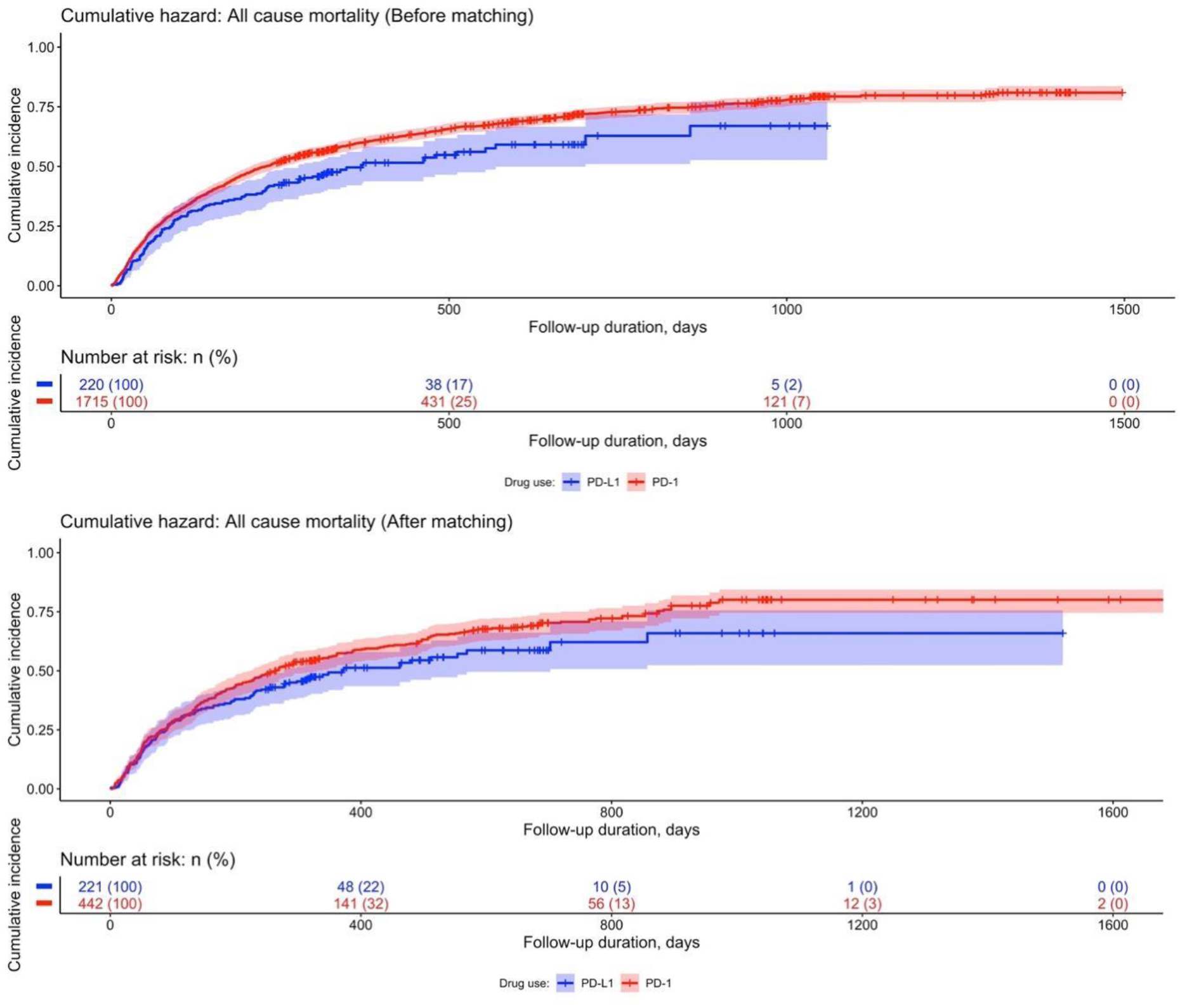
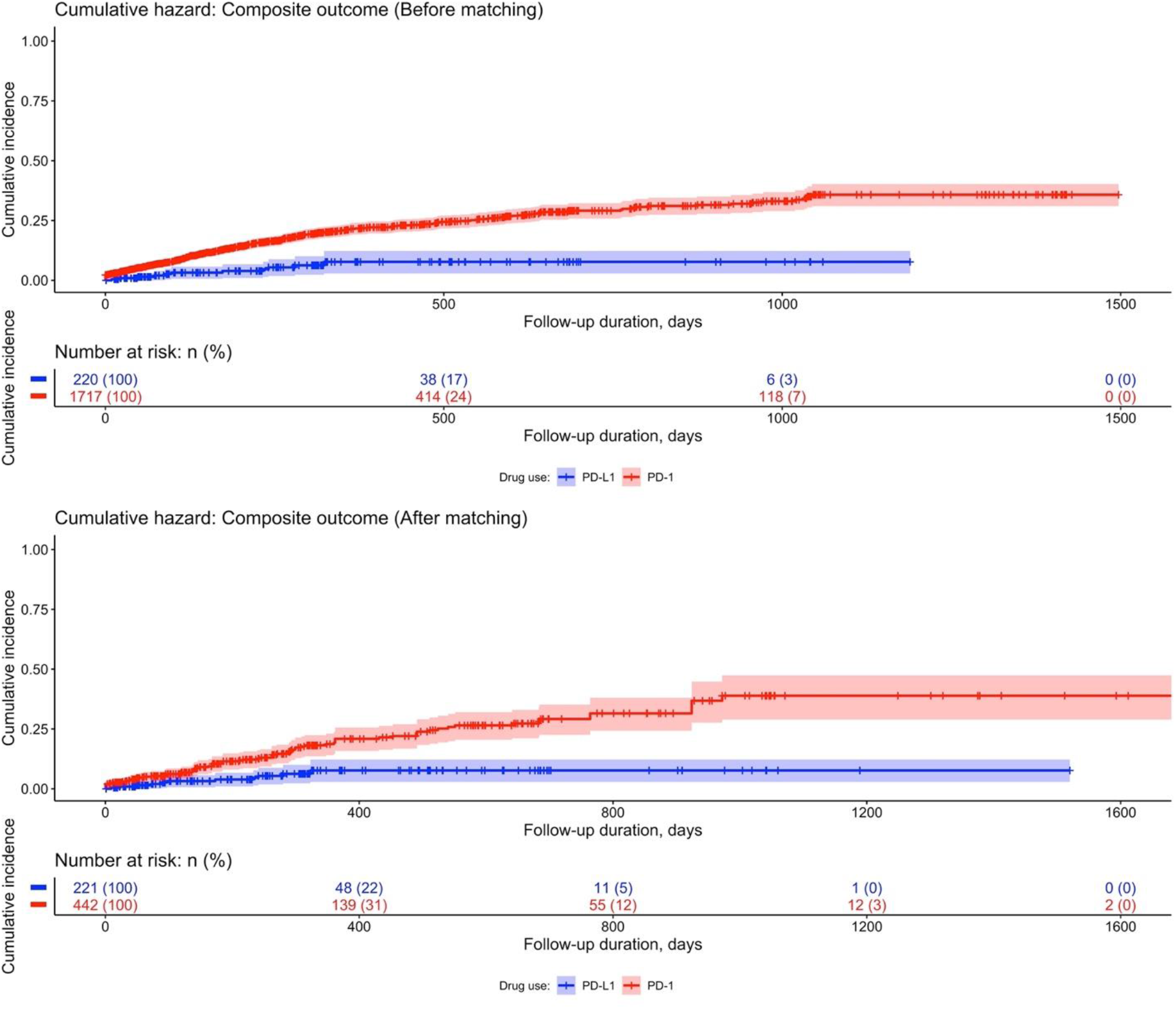
Cumulative incidence curves of new onset cardiac complications and all-cause mortality in cancer patients stratified by PD-1 or PD-L1 inhibitor use before and after 1:2 propensity score matching.

**Table 2.**
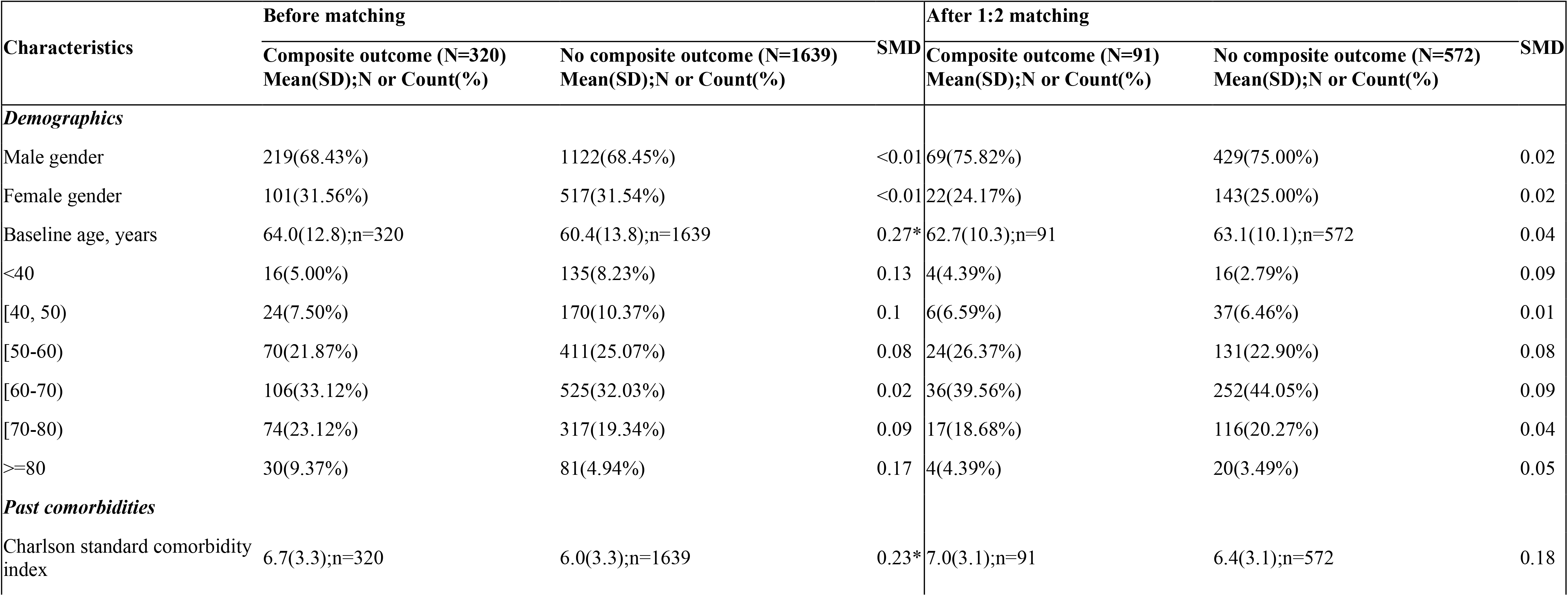
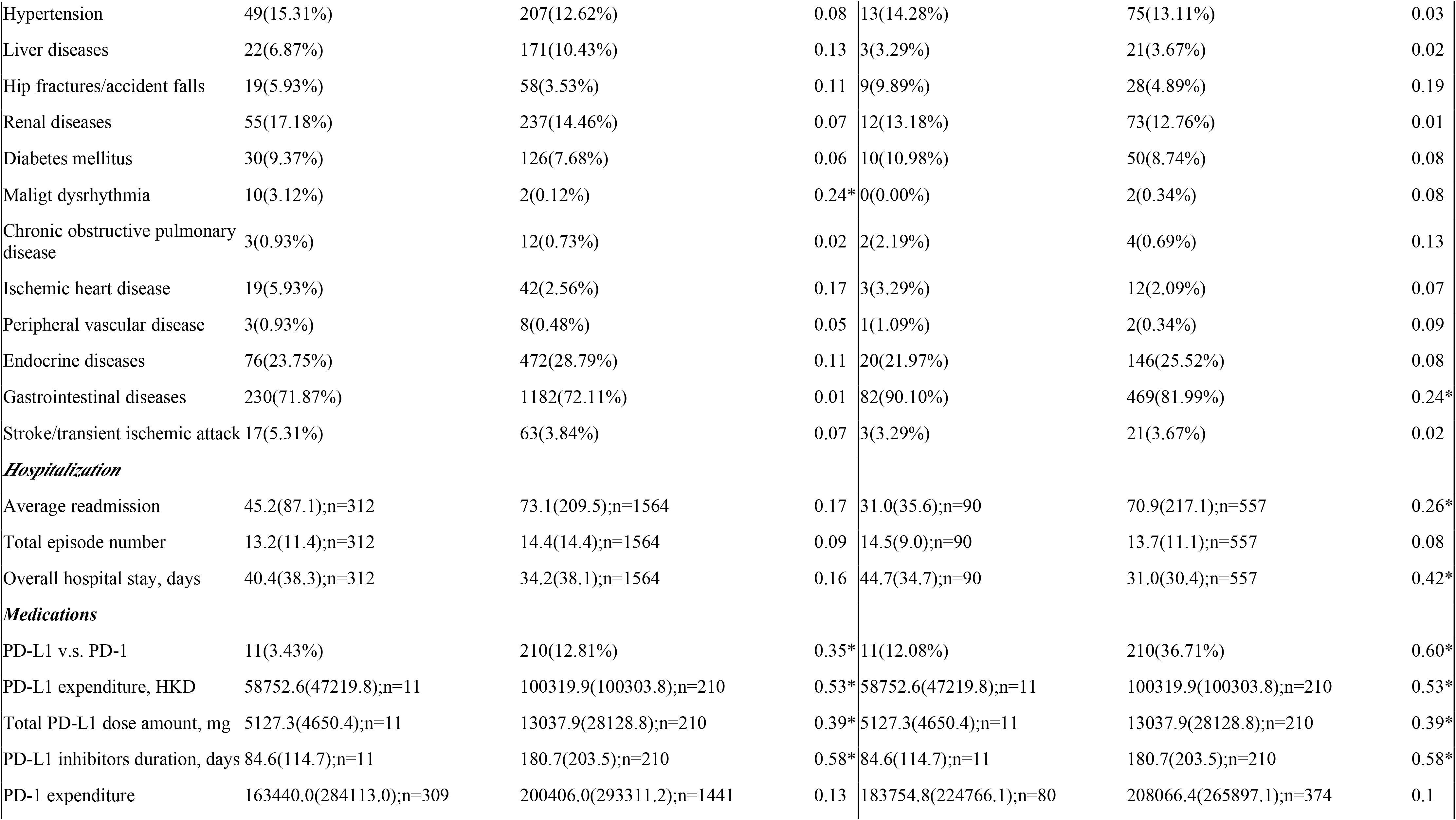
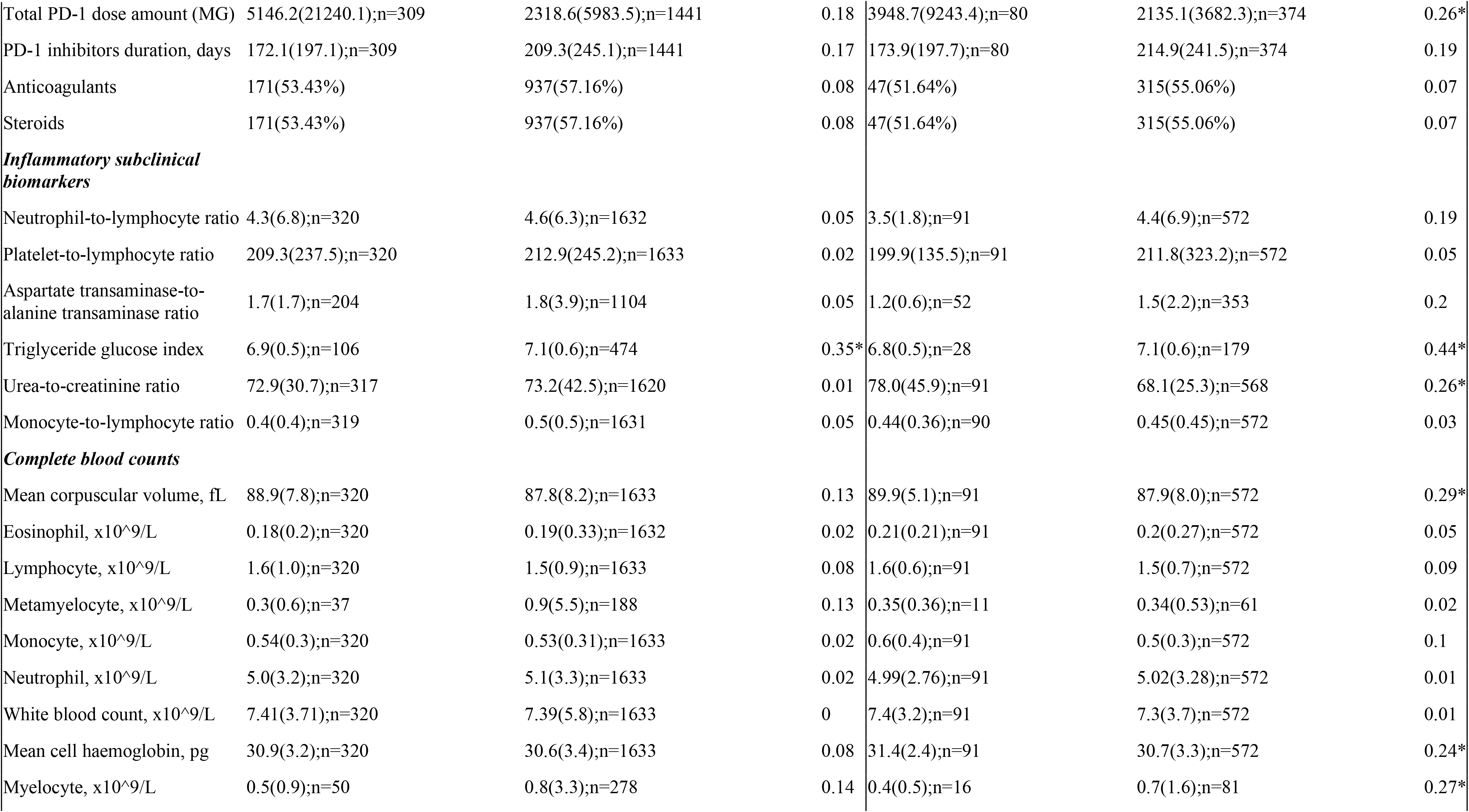
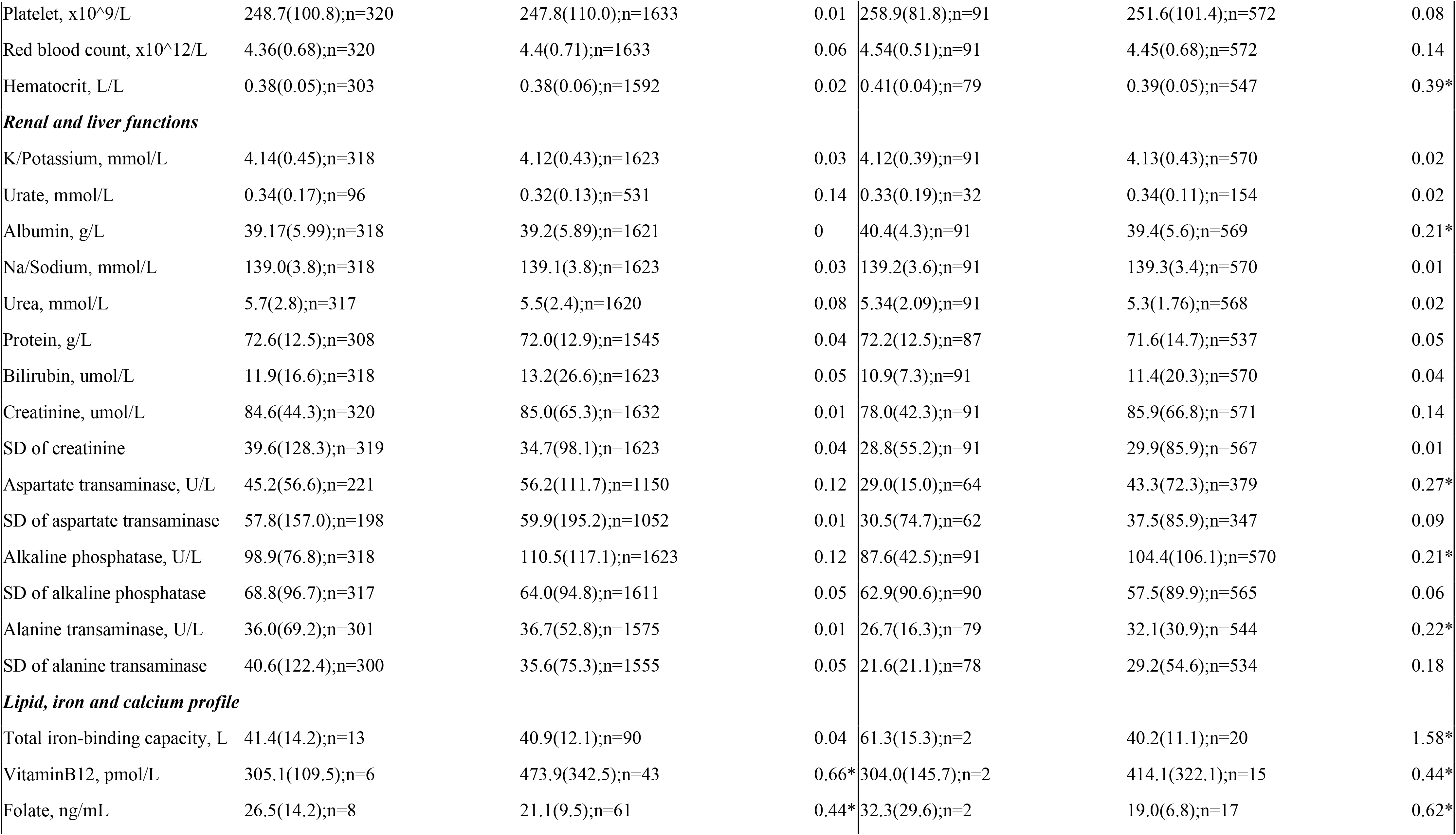
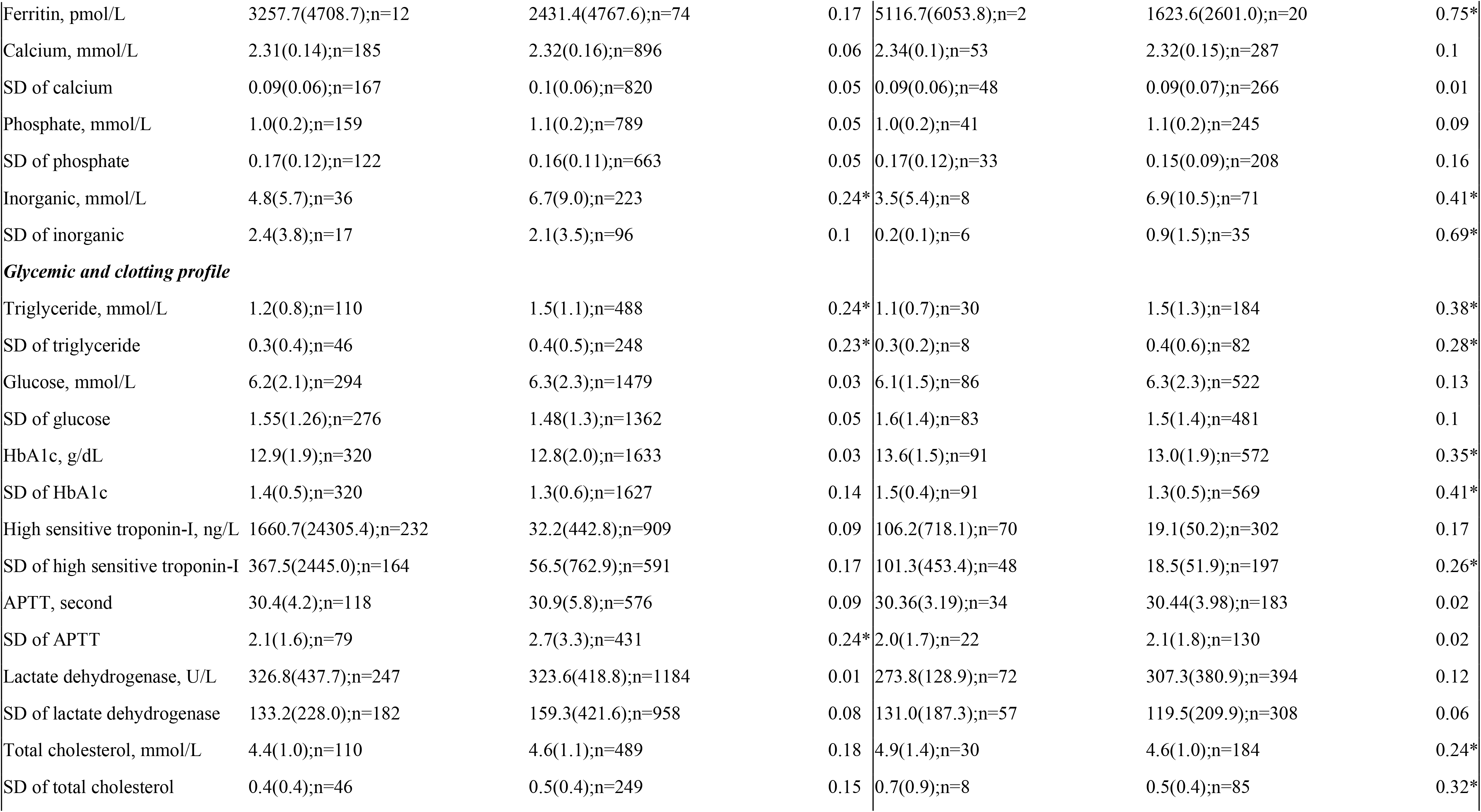
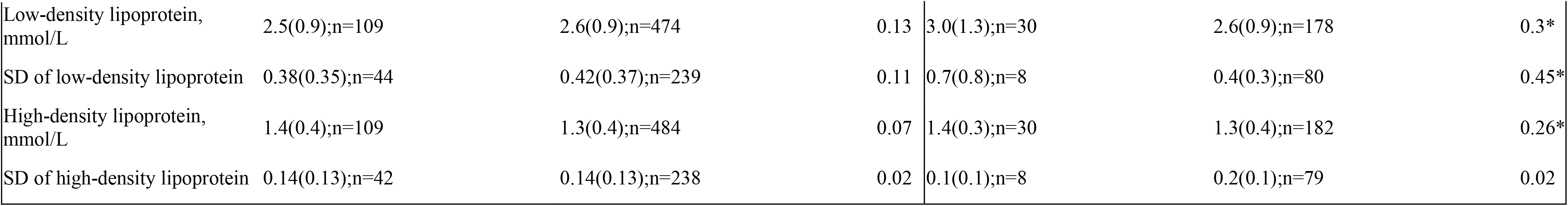
Clinical characteristics of patients who developed the composite outcome before and after 1:2 propensity score matching. * for *SMD* 0.2; # indicates the difference between patients with/without the composite outcome APTT: applied partial thromboplastin test; PD-1: Programmed death 1 inhibitors; PD-L1: programmed death 1 ligand inhibitors

| Characteristics | Before matching |  |  | After 1:2 matching |  |  |
| --- | --- | --- | --- | --- | --- | --- |
|  | Composite outcome (N=320)<br>Mean(SD);N or Count(%) | No composite outcome (N=1639)<br>Mean(SD);N or Count(%) | SMD | Composite outcome (N=91)<br>Mean(SD);N or Count(%) | No composite outcome (N=572)<br>Mean(SD);N or Count(%) | SMD |
| *Demographics* |  |  |  |  |  |  |
| Male gender | 219(68.43%) | 1122(68.45%) | <0.01 | 69(75.82%) | 429(75.00%) | 0.02 |
| Female gender | 101(31.56%) | 517(31.54%) | <0.01 | 22(24.17%) | 143(25.00%) | 0.02 |
| Baseline age, years | 64.0(12.8);n=320 | 60.4(13.8);n=1639 | 0.27* | 62.7(10.3);n=91 | 63.1(10.1);n=572 | 0.04 |
| <40 | 16(5.00%) | 135(8.23%) | 0.13 | 4(4.39%) | 16(2.79%) | 0.09 |
| [40, 50) | 24(7.50%) | 170(10.37%) | 0.1 | 6(6.59%) | 37(6.46%) | 0.01 |
| [50-60) | 70(21.87%) | 411(25.07%) | 0.08 | 24(26.37%) | 131(22.90%) | 0.08 |
| [60-70) | 106(33.12%) | 525(32.03%) | 0.02 | 36(39.56%) | 252(44.05%) | 0.09 |
| [70-80) | 74(23.12%) | 317(19.34%) | 0.09 | 17(18.68%) | 116(20.27%) | 0.04 |
| >=80 | 30(9.37%) | 81(4.94%) | 0.17 | 4(4.39%) | 20(3.49%) | 0.05 |
| *Past comorbidities* |  |  |  |  |  |  |
| Charlson standard comorbidity index | 6.7(3.3);n=320 | 6.0(3.3);n=1639 | 0.23* | 7.0(3.1);n=91 | 6.4(3.1);n=572 | 0.18 |
| Hypertension | 49(15.31%) | 207(12.62%) | 0.08 | 13(14.28%) | 75(13.11%) | 0.03 |
| Liver diseases | 22(6.87%) | 171(10.43%) | 0.13 | 3(3.29%) | 21(3.67%) | 0.02 |
| Hip fractures/accident falls | 19(5.93%) | 58(3.53%) | 0.11 | 9(9.89%) | 28(4.89%) | 0.19 |
| Renal diseases | 55(17.18%) | 237(14.46%) | 0.07 | 12(13.18%) | 73(12.76%) | 0.01 |
| Diabetes mellitus | 30(9.37%) | 126(7.68%) | 0.06 | 10(10.98%) | 50(8.74%) | 0.08 |
| Maligt dysrhythmia | 10(3.12%) | 2(0.12%) | 0.24* | 0(0.00%) | 2(0.34%) | 0.08 |
| Chronic obstructive pulmonary disease | 3(0.93%) | 12(0.73%) | 0.02 | 2(2.19%) | 4(0.69%) | 0.13 |
| Ischemic heart disease | 19(5.93%) | 42(2.56%) | 0.17 | 3(3.29%) | 12(2.09%) | 0.07 |
| Peripheral vascular disease | 3(0.93%) | 8(0.48%) | 0.05 | 1(1.09%) | 2(0.34%) | 0.09 |
| Endocrine diseases | 76(23.75%) | 472(28.79%) | 0.11 | 20(21.97%) | 146(25.52%) | 0.08 |
| Gastrointestinal diseases | 230(71.87%) | 1182(72.11%) | 0.01 | 82(90.10%) | 469(81.99%) | 0.24* |
| Stroke/transient ischemic attack | 17(5.31%) | 63(3.84%) | 0.07 | 3(3.29%) | 21(3.67%) | 0.02 |
| ***Hospitalization*** |  |  |  |  |  |  |
| Average readmission | 45.2(87.1);n=312 | 73.1(209.5);n=1564 | 0.17 | 31.0(35.6);n=90 | 70.9(217.1);n=557 | 0.26* |
| Total episode number | 13.2(11.4);n=312 | 14.4(14.4);n=1564 | 0.09 | 14.5(9.0);n=90 | 13.7(11.1);n=557 | 0.08 |
| Overall hospital stay, days | 40.4(38.3);n=312 | 34.2(38.1);n=1564 | 0.16 | 44.7(34.7);n=90 | 31.0(30.4);n=557 | 0.42* |
| ***Medications*** |  |  |  |  |  |  |
| PD-L1 v.s. PD-1 | 11(3.43%) | 210(12.81%) | 0.35* | 11(12.08%) | 210(36.71%) | 0.60* |
| PD-L1 expenditure, HKD | 58752.6(47219.8);n=11 | 100319.9(100303.8);n=210 | 0.53* | 58752.6(47219.8);n=11 | 100319.9(100303.8);n=210 | 0.53* |
| Total PD-L1 dose amount, mg | 5127.3(4650.4);n=11 | 13037.9(28128.8);n=210 | 0.39* | 5127.3(4650.4);n=11 | 13037.9(28128.8);n=210 | 0.39* |
| PD-L1 inhibitors duration, days | 84.6(114.7);n=11 | 180.7(203.5);n=210 | 0.58* | 84.6(114.7);n=11 | 180.7(203.5);n=210 | 0.58* |
| PD-1 expenditure | 163440.0(284113.0);n=309 | 200406.0(293311.2);n=1441 | 0.13 | 183754.8(224766.1);n=80 | 208066.4(265897.1);n=374 | 0.1 |
| Total PD-1 dose amount (MG) | 5146.2(21240.1);n=309 | 2318.6(5983.5);n=1441 | 0.18 | 3948.7(9243.4);n=80 | 2135.1(3682.3);n=374 | 0.26* |
| PD-1 inhibitors duration, days | 172.1(197.1);n=309 | 209.3(245.1);n=1441 | 0.17 | 173.9(197.7);n=80 | 214.9(241.5);n=374 | 0.19 |
| Anticoagulants | 171(53.43%) | 937(57.16%) | 0.08 | 47(51.64%) | 315(55.06%) | 0.07 |
| Steroids | 171(53.43%) | 937(57.16%) | 0.08 | 47(51.64%) | 315(55.06%) | 0.07 |
| *Inflammatory subclinical biomarkers* |  |  |  |  |  |  |
| Neutrophil-to-lymphocyte ratio | 4.3(6.8);n=320 | 4.6(6.3);n=1632 | 0.05 | 3.5(1.8);n=91 | 4.4(6.9);n=572 | 0.19 |
| Platelet-to-lymphocyte ratio | 209.3(237.5);n=320 | 212.9(245.2);n=1633 | 0.02 | 199.9(135.5);n=91 | 211.8(323.2);n=572 | 0.05 |
| Aspartate transaminase-to-alanine transaminase ratio | 1.7(1.7);n=204 | 1.8(3.9);n=1104 | 0.05 | 1.2(0.6);n=52 | 1.5(2.2);n=353 | 0.2 |
| Triglyceride glucose index | 6.9(0.5);n=106 | 7.1(0.6);n=474 | 0.35* | 6.8(0.5);n=28 | 7.1(0.6);n=179 | 0.44* |
| Urea-to-creatinine ratio | 72.9(30.7);n=317 | 73.2(42.5);n=1620 | 0.01 | 78.0(45.9);n=91 | 68.1(25.3);n=568 | 0.26* |
| Monocyte-to-lymphocyte ratio | 0.4(0.4);n=319 | 0.5(0.5);n=1631 | 0.05 | 0.44(0.36);n=90 | 0.45(0.45);n=572 | 0.03 |
| *Complete blood counts* |  |  |  |  |  |  |
| Mean corpuscular volume, fL | 88.9(7.8);n=320 | 87.8(8.2);n=1633 | 0.13 | 89.9(5.1);n=91 | 87.9(8.0);n=572 | 0.29* |
| Eosinophil, x10^9/L | 0.18(0.2);n=320 | 0.19(0.33);n=1632 | 0.02 | 0.21(0.21);n=91 | 0.2(0.27);n=572 | 0.05 |
| Lymphocyte, x10^9/L | 1.6(1.0);n=320 | 1.5(0.9);n=1633 | 0.08 | 1.6(0.6);n=91 | 1.5(0.7);n=572 | 0.09 |
| Metamyelocyte, x10^9/L | 0.3(0.6);n=37 | 0.9(5.5);n=188 | 0.13 | 0.35(0.36);n=11 | 0.34(0.53);n=61 | 0.02 |
| Monocyte, x10^9/L | 0.54(0.3);n=320 | 0.53(0.31);n=1633 | 0.02 | 0.6(0.4);n=91 | 0.5(0.3);n=572 | 0.1 |
| Neutrophil, x10^9/L | 5.0(3.2);n=320 | 5.1(3.3);n=1633 | 0.02 | 4.99(2.76);n=91 | 5.02(3.28);n=572 | 0.01 |
| White blood count, x10^9/L | 7.41(3.71);n=320 | 7.39(5.8);n=1633 | 0 | 7.4(3.2);n=91 | 7.3(3.7);n=572 | 0.01 |
| Mean cell haemoglobin, pg | 30.9(3.2);n=320 | 30.6(3.4);n=1633 | 0.08 | 31.4(2.4);n=91 | 30.7(3.3);n=572 | 0.24* |
| Myelocyte, x10^9/L | 0.5(0.9);n=50 | 0.8(3.3);n=278 | 0.14 | 0.4(0.5);n=16 | 0.7(1.6);n=81 | 0.27* |
| Platelet, x10^9/L | 248.7(100.8);n=320 | 247.8(110.0);n=1633 | 0.01 | 258.9(81.8);n=91 | 251.6(101.4);n=572 | 0.08 |
| Red blood count, x10^12/L | 4.36(0.68);n=320 | 4.4(0.71);n=1633 | 0.06 | 4.54(0.51);n=91 | 4.45(0.68);n=572 | 0.14 |
| Hematocrit, L/L | 0.38(0.05);n=303 | 0.38(0.06);n=1592 | 0.02 | 0.41(0.04);n=79 | 0.39(0.05);n=547 | 0.39* |
| ***Renal and liver functions*** |  |  |  |  |  |  |
| K/Potassium, mmol/L | 4.14(0.45);n=318 | 4.12(0.43);n=1623 | 0.03 | 4.12(0.39);n=91 | 4.13(0.43);n=570 | 0.02 |
| Urate, mmol/L | 0.34(0.17);n=96 | 0.32(0.13);n=531 | 0.14 | 0.33(0.19);n=32 | 0.34(0.11);n=154 | 0.02 |
| Albumin, g/L | 39.17(5.99);n=318 | 39.2(5.89);n=1621 | 0 | 40.4(4.3);n=91 | 39.4(5.6);n=569 | 0.21* |
| Na/Sodium, mmol/L | 139.0(3.8);n=318 | 139.1(3.8);n=1623 | 0.03 | 139.2(3.6);n=91 | 139.3(3.4);n=570 | 0.01 |
| Urea, mmol/L | 5.7(2.8);n=317 | 5.5(2.4);n=1620 | 0.08 | 5.34(2.09);n=91 | 5.3(1.76);n=568 | 0.02 |
| Protein, g/L | 72.6(12.5);n=308 | 72.0(12.9);n=1545 | 0.04 | 72.2(12.5);n=87 | 71.6(14.7);n=537 | 0.05 |
| Bilirubin, umol/L | 11.9(16.6);n=318 | 13.2(26.6);n=1623 | 0.05 | 10.9(7.3);n=91 | 11.4(20.3);n=570 | 0.04 |
| Creatinine, umol/L | 84.6(44.3);n=320 | 85.0(65.3);n=1632 | 0.01 | 78.0(42.3);n=91 | 85.9(66.8);n=571 | 0.14 |
| SD of creatinine | 39.6(128.3);n=319 | 34.7(98.1);n=1623 | 0.04 | 28.8(55.2);n=91 | 29.9(85.9);n=567 | 0.01 |
| Aspartate transaminase, U/L | 45.2(56.6);n=221 | 56.2(111.7);n=1150 | 0.12 | 29.0(15.0);n=64 | 43.3(72.3);n=379 | 0.27* |
| SD of aspartate transaminase | 57.8(157.0);n=198 | 59.9(195.2);n=1052 | 0.01 | 30.5(74.7);n=62 | 37.5(85.9);n=347 | 0.09 |
| Alkaline phosphatase, U/L | 98.9(76.8);n=318 | 110.5(117.1);n=1623 | 0.12 | 87.6(42.5);n=91 | 104.4(106.1);n=570 | 0.21* |
| SD of alkaline phosphatase | 68.8(96.7);n=317 | 64.0(94.8);n=1611 | 0.05 | 62.9(90.6);n=90 | 57.5(89.9);n=565 | 0.06 |
| Alanine transaminase, U/L | 36.0(69.2);n=301 | 36.7(52.8);n=1575 | 0.01 | 26.7(16.3);n=79 | 32.1(30.9);n=544 | 0.22* |
| SD of alanine transaminase | 40.6(122.4);n=300 | 35.6(75.3);n=1555 | 0.05 | 21.6(21.1);n=78 | 29.2(54.6);n=534 | 0.18 |
| ***Lipid, iron and calcium profile*** |  |  |  |  |  |  |
| Total iron-binding capacity, L | 41.4(14.2);n=13 | 40.9(12.1);n=90 | 0.04 | 61.3(15.3);n=2 | 40.2(11.1);n=20 | 1.58* |
| VitaminB12, pmol/L | 305.1(109.5);n=6 | 473.9(342.5);n=43 | 0.66* | 304.0(145.7);n=2 | 414.1(322.1);n=15 | 0.44* |
| Folate, ng/mL | 26.5(14.2);n=8 | 21.1(9.5);n=61 | 0.44* | 32.3(29.6);n=2 | 19.0(6.8);n=17 | 0.62* |
| Ferritin, pmol/L | 3257.7(4708.7);n=12 | 2431.4(4767.6);n=74 | 0.17 | 5116.7(6053.8);n=2 | 1623.6(2601.0);n=20 | 0.75* |
| Calcium, mmol/L | 2.31(0.14);n=185 | 2.32(0.16);n=896 | 0.06 | 2.34(0.1);n=53 | 2.32(0.15);n=287 | 0.1 |
| SD of calcium | 0.09(0.06);n=167 | 0.1(0.06);n=820 | 0.05 | 0.09(0.06);n=48 | 0.09(0.07);n=266 | 0.01 |
| Phosphate, mmol/L | 1.0(0.2);n=159 | 1.1(0.2);n=789 | 0.05 | 1.0(0.2);n=41 | 1.1(0.2);n=245 | 0.09 |
| SD of phosphate | 0.17(0.12);n=122 | 0.16(0.11);n=663 | 0.05 | 0.17(0.12);n=33 | 0.15(0.09);n=208 | 0.16 |
| Inorganic, mmol/L | 4.8(5.7);n=36 | 6.7(9.0);n=223 | 0.24* | 3.5(5.4);n=8 | 6.9(10.5);n=71 | 0.41* |
| SD of inorganic | 2.4(3.8);n=17 | 2.1(3.5);n=96 | 0.1 | 0.2(0.1);n=6 | 0.9(1.5);n=35 | 0.69* |
| *Glycemic and clotting profile* |  |  |  |  |  |  |
| Triglyceride, mmol/L | 1.2(0.8);n=110 | 1.5(1.1);n=488 | 0.24* | 1.1(0.7);n=30 | 1.5(1.3);n=184 | 0.38* |
| SD of triglyceride | 0.3(0.4);n=46 | 0.4(0.5);n=248 | 0.23* | 0.3(0.2);n=8 | 0.4(0.6);n=82 | 0.28* |
| Glucose, mmol/L | 6.2(2.1);n=294 | 6.3(2.3);n=1479 | 0.03 | 6.1(1.5);n=86 | 6.3(2.3);n=522 | 0.13 |
| SD of glucose | 1.55(1.26);n=276 | 1.48(1.3);n=1362 | 0.05 | 1.6(1.4);n=83 | 1.5(1.4);n=481 | 0.1 |
| HbA1c, g/dL | 12.9(1.9);n=320 | 12.8(2.0);n=1633 | 0.03 | 13.6(1.5);n=91 | 13.0(1.9);n=572 | 0.35* |
| SD of HbA1c | 1.4(0.5);n=320 | 1.3(0.6);n=1627 | 0.14 | 1.5(0.4);n=91 | 1.3(0.5);n=569 | 0.41* |
| High sensitive troponin-I, ng/L | 1660.7(24305.4);n=232 | 32.2(442.8);n=909 | 0.09 | 106.2(718.1);n=70 | 19.1(50.2);n=302 | 0.17 |
| SD of high sensitive troponin-I | 367.5(2445.0);n=164 | 56.5(762.9);n=591 | 0.17 | 101.3(453.4);n=48 | 18.5(51.9);n=197 | 0.26* |
| APTT, second | 30.4(4.2);n=118 | 30.9(5.8);n=576 | 0.09 | 30.36(3.19);n=34 | 30.44(3.98);n=183 | 0.02 |
| SD of APTT | 2.1(1.6);n=79 | 2.7(3.3);n=431 | 0.24* | 2.0(1.7);n=22 | 2.1(1.8);n=130 | 0.02 |
| Lactate dehydrogenase, U/L | 326.8(437.7);n=247 | 323.6(418.8);n=1184 | 0.01 | 273.8(128.9);n=72 | 307.3(380.9);n=394 | 0.12 |
| SD of lactate dehydrogenase | 133.2(228.0);n=182 | 159.3(421.6);n=958 | 0.08 | 131.0(187.3);n=57 | 119.5(209.9);n=308 | 0.06 |
| Total cholesterol, mmol/L | 4.4(1.0);n=110 | 4.6(1.1);n=489 | 0.18 | 4.9(1.4);n=30 | 4.6(1.0);n=184 | 0.24* |
| SD of total cholesterol | 0.4(0.4);n=46 | 0.5(0.4);n=249 | 0.15 | 0.7(0.9);n=8 | 0.5(0.4);n=85 | 0.32* |
| Low-density lipoprotein,<br>mmol/L | 2.5(0.9);n=109 | 2.6(0.9);n=474 | 0.13 | 3.0(1.3);n=30 | 2.6(0.9);n=178 | 0.3* |
| SD of low-density lipoprotein | 0.38(0.35);n=44 | 0.42(0.37);n=239 | 0.11 | 0.7(0.8);n=8 | 0.4(0.3);n=80 | 0.45* |
| High-density lipoprotein,<br>mmol/L | 1.4(0.4);n=109 | 1.3(0.4);n=484 | 0.07 | 1.4(0.3);n=30 | 1.3(0.4);n=182 | 0.26* |
| SD of high-density lipoprotein | 0.14(0.13);n=42 | 0.14(0.13);n=238 | 0.02 | 0.1(0.1);n=8 | 0.2(0.1);n=79 | 0.02 |

**Table 3.** Significant univariate predictors of new onset cardiac complication outcome and all-cause mortality before and after 1:2 propensity score matching. * for p≤ 0.05, ** for p ≤ 0.01, *** for p ≤ 0.001; APTT: applied partial thromboplastin test; PD-1: Programmed death 1 inhibitors; PD-L1: programmed death 1 ligand inhibitors; CI: Confidence interval; HR: Hazard ratio.

| Characteristics | Before matching |  | After 1:2 matching |  |
| --- | --- | --- | --- | --- |
|  | All-cause mortality | Composite outcome | All-cause mortality | Composite outcome |
|  | HR [95% CI];P value | HR [95% CI];P value | HR [95% CI];P value | HR [95% CI];P value |
| *Demographics* |  |  |  |  |
| Male gender | 0.98[0.87-1.10];0.6878 | 1.00[0.78-1.29];0.9898 | 1.03[0.83-1.29];0.7733 | 1.12[0.67-1.88];0.6546 |
| Female gender | 1.0[Reference] | 1.0[Reference] | 1.0[Reference] | 1.0[Reference] |
| Baseline age, years | 1.00[1.00-1.01];0.1533 | 1.01[1.01-1.02];0.0016** | 1.01[1.00-1.01];0.2976 | 1.00[0.98-1.02];0.9421 |
| <40 | 1.0[Reference] | 1.0[Reference] | 1.0[Reference] | 1.0[Reference] |
| [40, 50) | 1.02[0.85-1.22];0.8400 | 0.72[0.46-1.12];0.1450 | 1.07[0.73-1.56];0.7308 | 0.96[0.39-2.38];0.9321 |
| [50-60) | 1.04[0.92-1.18];0.5569 | 0.97[0.74-1.28];0.8386 | 0.87[0.69-1.09];0.2345 | 1.20[0.74-1.95];0.4528 |
| [60-70) | 1.02[0.91-1.14];0.7455 | 1.05[0.82-1.35];0.6880 | 0.98[0.81-1.19];0.8684 | 0.71[0.45-1.11];0.1309 |
| [70-80) | 0.98[0.85-1.12];0.7150 | 1.19[0.91-1.57];0.2103 | 1.16[0.92-1.47];0.2172 | 1.13[0.66-1.96];0.6528 |
| >=80 | 1.07[0.86-1.35];0.5366 | 1.40[0.89-2.21];0.1436 | 1.03[0.63-1.70];0.9074 | 1.51[0.55-4.13];0.4210 |
| *Past comorbidities* |  |  |  |  |
| Charlson standard comorbidity index | 1.05[1.04-1.07];<0.0001*** | 1.08[1.04-1.12];<0.0001*** | 1.08[1.04-1.11];<0.0001*** | 1.07[1.00-1.15];0.0589 |
| Hypertension | 1.20[1.02-1.40];0.0262* | 1.11[0.78-1.59];0.5645 | 1.32[1.00-1.73];0.0481* | 1.18[0.61-2.29];0.6236 |
| Liver diseases | 1.24[1.04-1.47];0.0163* | 0.77[0.49-1.22];0.2689 | 1.46[0.91-2.35];0.1148 | 1.32[0.42-4.18];0.6381 |
| Hip fractures/accident falls | 1.26[0.96-1.64];0.0910 | 1.98[1.21-3.23];0.0065** | 0.79[0.50-1.25];0.3174 | 2.21[1.10-4.41];0.0253* |
| Renal diseases | 1.02[0.88-1.19];0.7670 | 1.06[0.77-1.48];0.7155 | 1.05[0.79-1.39];0.7571 | 1.24[0.67-2.29];0.4871 |
| Diabetes mellitus | 1.13[0.92-1.37];0.2369 | 1.05[0.67-1.66];0.8204 | 1.12[0.80-1.57];0.4963 | 1.12[0.52-2.43];0.7733 |
| Maligt dysrhythmia | 2.09[1.12-3.89];0.0206* | 11.11[5.88-20.98];<0.0001*** | 0.00[0.00-Inf];0.9877 | 0.00[0.00-Inf];0.9946 |
| Chronic obstructive pulmonary disease | 1.42[0.78-2.56];0.2515 | 1.83[0.59-5.72];0.2971 | 2.04[0.84-4.94];0.1133 | 5.51[1.34-22.64];0.0179* |
| Ischemic heart disease | 0.98[0.72-1.34];0.8984 | 1.30[0.69-2.44];0.4160 | 0.98[0.49-1.98];0.9639 | 0.70[0.10-5.07];0.7271 |
| Peripheral vascular disease | 1.10[0.57-2.12];0.7774 | 1.93[0.62-6.03];0.2570 | 0.87[0.22-3.51];0.8502 | 2.51[0.35-18.07];0.3611 |
| Endocrine diseases | 1.04[0.93-1.18];0.4907 | 0.76[0.58-1.00];0.0538 | 1.04[0.84-1.29];0.7304 | 0.75[0.44-1.28];0.2934 |
| Gastrointestinal diseases | 1.03[0.92-1.17];0.5810 | 0.94[0.73-1.21];0.6327 | 1.22[0.93-1.59];0.1534 | 1.71[0.85-3.41];0.1296 |
| Stroke/transient ischemic attack | 1.11[0.85-1.46];0.4279 | 1.02[0.54-1.92];0.9477 | 1.48[0.91-2.40];0.1149 | 0.96[0.24-3.92];0.9586 |
| ***Hospitalization*** |  |  |  |  |
| Average readmission | 1.000[1.000-1.000];0.2724 | 0.998[0.997-1.000];0.0625 | 1.000[1.000-1.001];0.0823 | 0.99[0.98-1.00];0.0080** |
| Total episode number | 0.95[0.95-0.96];<0.0001*** | 0.96[0.95-0.97];<0.0001*** | 0.96[0.95-0.97];<0.0001*** | 0.97[0.95-0.99];0.0050** |
| Overall hospital stay, days | 1.001[0.999-1.002];0.3779 | 1.001[0.999-1.004];0.3117 | 1.00[1.00-1.01];0.0004*** | 1.01[1.00-1.01];0.0040** |
| ***Medications*** |  |  |  |  |
| PD-L1 v.s. PD-1 | 0.77[0.64-0.93];0.0078** | 0.32[0.18-0.59];0.0002*** | 0.80[0.65-1.00];0.0463* | 0.34[0.18-0.65];0.0010** |
| PD-L1 expenditure, HKD | 1.000[1.000-1.000];<0.0001*** | 1.000[1.000-1.000];0.0528 | 1.000[1.000-1.000];<0.0001*** | 1.000[1.000-1.000];0.0528 |
| Total PD-L1 dose amount, mg | 1.000[1.000-1.000];0.0703 | 1.000[1.000-1.000];0.2410 | 1.000[1.000-1.000];0.0703 | 1.000[1.000-1.000];0.2410 |
| PD-L1 inhibitors duration, days | 1.00[0.99-1.00];<0.0001*** | 0.99[0.99-1.00];0.0263* | 1.00[0.99-1.00];<0.0001*** | 0.99[0.99-1.00];0.0263* |
| PD-1 expenditure | 1.000[1.000-1.000];<0.0001*** | 1.000[1.000-1.000];<0.0001*** | 1.000[1.000-1.000];<0.0001*** | 1.000[1.000-1.001];0.0018** |
| Total PD-1 dose amount (MG) | 1.000[1.000-1.000];<0.0001*** | 1.000[1.000-1.000];0.1842 | 1.000[1.000-1.000];0.0008*** | 1.000[1.000-1.000];0.0958 |
| PD-1 inhibitors duration, days | 0.997[0.997-0.998];<0.0001*** | 0.998[0.997-0.998];<0.0001*** | 0.998[0.997-0.998];<0.0001*** | 0.998[0.997-0.999];0.0007*** |
| Anticoagulants | 0.81[0.73-0.91];0.0002*** | 0.73[0.58-0.93];0.0097** | 0.89[0.74-1.08];0.2423 | 0.78[0.50-1.20];0.2574 |
| Steroids | 0.81[0.73-0.91];0.0002*** | 0.73[0.58-0.93];0.0097** | 0.89[0.74-1.08];0.2423 | 0.78[0.50-1.20];0.2574 |
| ***Inflammatory subclinical biomarkers*** |  |  |  |  |
| Neutrophil-to-lymphocyte ratio | 1.01[1.00-1.01];0.0296* | 1.00[0.98-1.02];0.8231 | 1.01[1.00-1.02];0.1025 | 0.97[0.92-1.03];0.3646 |
| Platelet-to-lymphocyte ratio | 1.000[1.000-1.000];0.0261* | 1.000[1.000-1.001];0.5907 | 1.000[1.000-1.000];0.1087 | 1.000[0.999-1.001];0.9257 |
| Aspartate transaminase-to-alanine transaminase ratio | 1.03[1.01-1.04];<0.0001*** | 1.01[0.97-1.06];0.6492 | 1.09[1.04-1.14];0.0004*** | 0.96[0.71-1.29];0.7683 |
| Triglyceride glucose index | 0.97[0.83-1.13];0.6770 | 0.59[0.41-0.85];0.0040** | 0.85[0.66-1.09];0.2061 | 0.55[0.30-1.02];0.0594 |
| Urea-to-creatinine ratio | 1.000[0.999-1.001];0.9657 | 1.000[0.997-1.002];0.9081 | 1.00[1.00-1.01];0.2003 | 1.01[1.00-1.01];0.0007*** |
| Monocyte-to-lymphocyte ratio | 1.07[0.99-1.16];0.0998 | 0.99[0.79-1.23];0.9303 | 1.20[1.02-1.43];0.0305* | 1.07[0.69-1.67];0.7559 |
| ***Complete blood counts*** |  |  |  |  |
| Mean corpuscular volume, fL | 1.00[0.99-1.01];0.9942 | 1.01[1.00-1.03];0.1496 | 1.00[0.98-1.01];0.5845 | 1.03[1.00-1.07];0.0470* |
| Eosinophil, x10^9/L | 0.74[0.58-0.93];0.0105* | 0.84[0.54-1.31];0.4333 | 0.81[0.53-1.23];0.3143 | 1.14[0.49-2.64];0.7632 |
| Lymphocyte, x10^9/L | 0.98[0.92-1.05];0.6221 | 1.08[0.97-1.20];0.1514 | 0.96[0.83-1.10];0.5296 | 1.07[0.78-1.47];0.6699 |
| Metamyelocyte, x10^9/L | 0.87[0.70-1.07];0.1830 | 0.78[0.46-1.31];0.3392 | 0.71[0.40-1.28];0.2549 | 0.74[0.21-2.63];0.6383 |
| Monocyte, x10^9/L | 1.24[1.06-1.45];0.0078** | 1.23[0.87-1.73];0.2362 | 1.51[1.14-2.00];0.0045** | 1.65[0.88-3.11];0.1194 |
| Neutrophil, x10^9/L | 1.02[1.01-1.04];0.0013** | 1.01[0.98-1.05];0.4025 | 1.03[1.01-1.06];0.0129* | 1.02[0.96-1.08];0.5817 |
| White blood count, x10^9/L | 1.02[1.01-1.03];<0.0001*** | 1.02[1.00-1.05];0.1069 | 1.03[1.00-1.05];0.0187* | 1.02[0.97-1.08];0.4938 |
| Mean cell haemoglobin, pg | 0.99[0.97-1.01];0.2267 | 1.01[0.97-1.05];0.6190 | 0.96[0.94-0.99];0.0133* | 1.05[0.97-1.14];0.1981 |
| Myelocyte, x10^9/L | 0.93[0.84-1.02];0.1131 | 0.88[0.66-1.18];0.3893 | 0.83[0.66-1.04];0.1098 | 0.83[0.50-1.39];0.4823 |
| Platelet, x10^9/L | 1.001[1.000-1.001];0.0388* | 1.001[1.000-1.002];0.2819 | 1.002[1.001-1.003];0.0015** | 1.002[0.999-1.004];0.1452 |
| Red blood count, x10^12/L | 0.93[0.86-1.01];0.0818 | 0.93[0.78-1.10];0.3688 | 0.91[0.79-1.06];0.2409 | 1.26[0.89-1.78];0.1925 |
| Hematocrit, L/L | 0.30[0.11-0.80];0.0163* | 0.75[0.09-6.67];0.7997 | 0.18[0.03-1.13];0.0671 | 566.25[4.88-65751.46];0.0090** |
| ***Renal and liver functions*** |  |  |  |  |
| K/Potassium, mmol/L | 0.91[0.80-1.03];0.1211 | 1.03[0.79-1.35];0.8120 | 0.87[0.69-1.09];0.2157 | 0.92[0.56-1.52];0.7440 |
| Urate, mmol/L | 1.54[0.72-3.30];0.2671 | 5.04[0.95-26.62];0.0570 | 1.71[0.32-9.18];0.5314 | 0.65[0.02-19.52];0.8040 |
| Albumin, g/L | 0.97[0.96-0.98];<0.0001*** | 0.98[0.96-1.00];0.0540 | 0.98[0.96-0.99];0.0061** | 1.04[0.99-1.09];0.1011 |
| Na/Sodium, mmol/L | 0.97[0.96-0.98];<0.0001*** | 0.97[0.95-1.00];0.0227* | 0.95[0.92-0.97];0.0001*** | 0.97[0.91-1.05];0.4786 |
| Urea, mmol/L | 0.99[0.97-1.02];0.5540 | 1.02[0.98-1.06];0.4164 | 0.98[0.93-1.03];0.4672 | 1.00[0.89-1.13];0.9939 |
| Protein, g/L | 0.99[0.99-1.00];0.0001*** | 1.00[0.99-1.01];0.8901 | 0.99[0.99-1.00];0.0087** | 1.00[0.98-1.02];0.9227 |
| Bilirubin, umol/L | 1.003[1.002-1.005];<0.0001*** | 1.00[1.00-1.01];0.5730 | 1.00[1.00-1.01];0.9330 | 1.00[0.99-1.01];0.9730 |
| Creatinine, umol/L | 1.001[1.000-1.001];0.2210 | 1.000[0.998-1.002];0.7707 | 1.001[0.999-1.002];0.2969 | 1.00[0.99-1.00];0.4195 |
| SD of creatinine | 1.001[1.000-1.001];0.0001*** | 1.001[1.000-1.002];0.0719 | 1.001[1.000-1.001];0.0974 | 1.000[0.998-1.002];0.7760 |
| Aspartate transaminase, U/L | 1.001[1.001-1.001];<0.0001*** | 1.000[0.998-1.002];0.8846 | 1.001[0.999-1.002];0.4383 | 0.99[0.98-1.01];0.2798 |
| SD of aspartate transaminase | 1.001[1.000-1.001];<0.0001*** | 1.000[1.000-1.001];0.2117 | 1.001[1.000-1.002];0.0155* | 1.001[0.997-1.004];0.6500 |
| Alkaline phosphatase, U/L | 1.001[1.001-1.002];<0.0001*** | 1.000[0.999-1.002];0.7097 | 1.000[0.999-1.001];0.4551 | 1.00[0.99-1.00];0.2454 |
| SD of alkaline phosphatase | 1.002[1.002-1.003];<0.0001*** | 1.002[1.001-1.003];0.0014** | 1.003[1.002-1.003];<0.0001*** | 1.002[1.000-1.004];0.0359* |
| Alanine transaminase, U/L | 1.000[1.000-1.001];0.3737 | 1.000[0.998-1.002];0.8684 | 0.999[0.995-1.002];0.3897 | 0.99[0.98-1.00];0.0914 |
| SD of alanine transaminase | 1.001[1.001-1.002];<0.0001*** | 1.001[1.000-1.002];0.0177* | 1.001[1.000-1.003];0.0741 | 1.00[0.99-1.00];0.5347 |
| ***Lipid, iron and calcium profile*** |  |  |  |  |
| Total iron-binding capacity, L | 1.00[0.98-1.02];0.9971 | 1.00[0.95-1.05];0.9678 | 0.99[0.95-1.04];0.7518 | 3.87[0.00-Inf];0.9993 |
| VitaminB12, pmol/L | 0.999[0.998-1.000];0.1739 | 1.00[0.99-1.00];0.1813 | 1.000[0.998-1.002];0.7853 | 1.00[0.99-1.01];0.8806 |
| Folate, ng/mL | 1.00[0.97-1.03];0.9877 | 1.04[0.97-1.12];0.2752 | 1.00[0.96-1.05];0.8835 | 1.04[0.95-1.14];0.3791 |
| Ferritin, pmol/L | 1.000[1.000-1.000];0.4438 | 1.000[1.000-1.000];0.3647 | 1.000[1.000-1.000];0.2525 | 1.000[1.000-1.001];0.1003 |
| Calcium, mmol/L | 0.64[0.39-1.06];0.0826 | 0.41[0.13-1.26];0.1212 | 0.91[0.36-2.28];0.8345 | 1.40[0.20-9.84];0.7350 |
| SD of calcium | 12.70[5.21-30.97];<0.0001*** | 2.10[0.13-34.34];0.6015 | 4.69[1.00-22.02];0.0499* | 2.32[0.04-132.03];0.6840 |
| Phosphate, mmol/L | 0.83[0.59-1.18];0.3059 | 0.79[0.36-1.74];0.5548 | 0.98[0.50-1.90];0.9464 | 0.61[0.14-2.69];0.5115 |
| SD of phosphate | 5.97[3.23-11.04];<0.0001*** | 7.44[1.81-30.57];0.0054** | 3.91[0.74-20.76];0.1098 | 18.22[0.41-815.46];0.1345 |
| Inorganic, mmol/L | 1.01[1.00-1.03];0.0836 | 0.99[0.94-1.04];0.7433 | 1.04[1.01-1.08];0.0055** | 0.97[0.84-1.13];0.7058 |
| SD of inorganic | 1.00[0.95-1.07];0.8984 | 0.98[0.82-1.17];0.8125 | 1.08[0.85-1.36];0.5374 | 0.09[0.00-21.93];0.3871 |
| ***Glycemic and clotting profile*** |  |  |  |  |
| Triglyceride, mmol/L | 0.96[0.88-1.05];0.3264 | 0.74[0.56-0.99];0.0436* | 0.90[0.78-1.04];0.1556 | 0.61[0.35-1.09];0.0933 |
| SD of triglyceride | 0.84[0.64-1.11];0.2196 | 0.20[0.04-1.02];0.0526 | 0.69[0.41-1.17];0.1655 | 0.54[0.08-3.53];0.5213 |
| Glucose, mmol/L | 1.02[0.99-1.04];0.1738 | 1.00[0.94-1.05];0.9179 | 0.99[0.95-1.03];0.5861 | 0.90[0.79-1.04];0.1558 |
| SD of glucose | 1.06[1.02-1.11];0.0032** | 1.07[0.98-1.16];0.1337 | 1.07[1.00-1.14];0.0580 | 1.07[0.92-1.23];0.3816 |
| HbA1c, g/dL | 0.97[0.95-1.00];0.0539 | 1.00[0.94-1.06];0.9969 | 0.95[0.91-1.00];0.0543 | 1.19[1.05-1.34];0.0072** |
| SD of HbA1c | 1.58[1.44-1.74];<0.0001*** | 1.51[1.23-1.85];0.0001*** | 1.52[1.29-1.79];<0.0001*** | 1.94[1.36-2.76];0.0003*** |
| High sensitive troponin-I, ng/L | 1.000[1.000-1.000];0.9239 | 1.000[1.000-1.000];0.1181 | 1.000[1.000-1.000];0.9382 | 1.000[1.000-1.001];0.1785 |
| SD of high sensitive troponin-I | 1.000[1.000-1.000];0.7182 | 1.000[1.000-1.000];0.0044** | 1.000[0.999-1.001];0.9978 | 1.000[1.000-1.001];0.2563 |
| APTT, second | 1.01[1.00-1.03];0.1426 | 0.98[0.93-1.03];0.3733 | 0.99[0.95-1.03];0.6325 | 0.98[0.89-1.08];0.6750 |
| SD of APTT | 1.05[1.02-1.08];0.0006*** | 0.99[0.90-1.10];0.8880 | 1.01[0.90-1.13];0.8927 | 1.10[0.86-1.42];0.4432 |
| Lactate dehydrogenase, U/L | 1.000[1.000-1.000];<0.0001*** | 1.000[1.000-1.001];0.0165* | 1.000[1.000-1.001];0.0001*** | 1.000[0.999-1.001];0.6208 |
| SD of lactate dehydrogenase | 1.000[1.000-1.000];<0.0001*** | 1.000[1.000-1.001];0.0229* | 1.002[1.001-1.002];<0.0001*** | 1.002[1.000-1.003];0.0056** |
| Total cholesterol, mmol/L | 0.98[0.90-1.07];0.6769 | 0.92[0.75-1.13];0.4335 | 1.05[0.90-1.22];0.5598 | 1.42[1.00-2.00];0.0490* |
| SD of total cholesterol | 1.14[0.84-1.55];0.3905 | 0.93[0.42-2.07];0.8677 | 1.23[0.73-2.07];0.4394 | 2.73[0.81-9.19];0.1058 |
| Low-density lipoprotein, mmol/L | 1.00[0.90-1.11];0.9905 | 0.96[0.76-1.22];0.7514 | 1.14[0.96-1.37];0.1398 | 1.67[1.14-2.45];0.0091** |
| SD of low-density lipoprotein | 1.22[0.84-1.78];0.2952 | 1.13[0.45-2.83];0.7919 | 1.66[0.77-3.55];0.1928 | 7.64[1.84-31.77];0.0051** |
| High-density lipoprotein, mmol/L | 0.93[0.73-1.19];0.5774 | 1.27[0.77-2.11];0.3530 | 0.97[0.62-1.50];0.8772 | 1.96[0.82-4.71];0.1309 |
| SD of high-density lipoprotein | 2.42[0.85-6.87];0.0971 | 1.55[0.11-21.84];0.7461 | 1.32[0.19-9.12];0.7758 | 1.09[0.01-164.28];0.9719 |

Univariate Cox regression identified significant predictors of the primary composite outcome and all-cause mortality before and after propensity score matching (Table 3). Compared with PD-1 inhibitor treatment, PD-L1 inhibitor treatment was significantly associated with a lower risk of composite outcome both before (hazard ratio [HR]: 0.32, 95% CI: [0.18-0.59], P value=0.0002***) and after matching (HR: 0.34, 95% CI: [0.18-0.65], P value=0.001**), and lower all-cause mortality risk before matching (HR: 0.77, 95% CI: [0.64-0.93], P value=0.0078**) and after matching (HR: 0.80, 95% CI: [0.65-1.00], P value=0.0463).

More PD-L1 expenditure (HR:1.000; 95% CI: 1.000-1.000; P value=0.0528), shorter PD-L1 inhibitors duration (HR:0.99; 95% CI: 0.99-1.00; P value=0.0263*), more PD-1 expenditure (HR:1.000; 95% CI: 1.000-1.001; P value=0.0018**), and shorter PD-1 inhibitors duration (HR:0.998; 95% CI: 0.997-0.999; P value=0.0007***) were associated with new onset cardiac composite outcome in the matched cohort. Significant laboratory examinations significantly associated with new onset cardiac composite outcome include higher levels of mean corpuscular volume (HR:1.03; 95% CI: 1.00-1.07; P value=0.0470*), hematocrit (HR:566.25; 95% CI: 4.88-65751.46; P value=0.0090**), HbA1c (HR:1.19; 95% CI: 1.05-1.34; P value=0.0072**), total cholesterol (HR:1.42; 95% CI: 1.00-2.00; P value=0.0490*), and low-density lipoprotein (HR:1.67; 95% CI: 1.14-2.45; P value=0.0091**).

In addition, greater variability in laboratory tests, including the standard deviations (SD) of alkaline phosphatase (HR:1.002; 95% CI: 1.000-1.004; P value=0.0359*), HbA1c (HR:1.94; 95% CI: 1.36-2.76; P value=0.0003***), lactate dehydrogenase (HR:1.002; 95% CI: 1.000-1.003; P value=0.0056**), and low-density lipoprotein (HR:7.64; 95% CI: 1.84-31.77; P value=0.0051**) were significantly associated with the composite outcome. The boxplots of significant measures of variability stratified by PD-1/PD-L1 inhibitor treatment in the matched cohort are shown in Figure 5.

### Healthcare utilization before and after treatment with PD-1/PD-L1 inhibitors

Longer overall cumulative hospital stay (HR:1.01; 95% CI: 1.00-1.01; P value=0.0040**) and longer hospital stay after PD-1/PD-L1 drug use (HR:1.01; 95% CI: 1.00-1.01; P value=0.0040**) were significantly associated with the composite outcome. Furthermore, hospitalization characteristics before and after PD-1/PD-L1 treatment were compared in the subset of patients who developed the adverse outcomes, in both the unmatched and matched cohorts (Table 4). Patients who developed cardiovascular complications had a shorter average readmission interval, a higher number of hospitalizations and a longer duration of hospital stay after PD-1/PD-L1 treatment (P<0.0001).

**Table 4.** Comparisons of hospitalization characteristics before and after PD1/PD-L1 treatment in patients with new onset heart failure, acute myocardial infarction, atrial fibrillation, and atrial flutter. * for p≤ 0.05, ** for p ≤ 0.01, *** for p ≤ 0.001; PD-1: Programmed death 1 inhibitors; PD-L1: programmed death 1 ligand inhibitors.

| Before matching |  |  |  | After 1:1 matching |  |  |
| --- | --- | --- | --- | --- | --- | --- |
| Hospitalization characteristics | Before treatment Mean(SD);N or Count(%) | After treatment Mean(SD);N or Count(%) | P value | Before treatment Mean(SD);N or Count(%) | After treatment Mean(SD);N or Count(%) | P value |
| *Heart failure (N=244)* |  |  |  | *Heart failure (N=75)* |  |  |
| Average readmission interval between episodes, days | 299.3(680.4);n=202 | 45.5(95.9);n=244 | <0.0001*** | 358.6(922.4);n=61 | 38.1(45.2);n=75 | <0.0001*** |
| No. of episodes | 8.8(7.4);n=202 | 13.8(12.3);n=244 | <0.0001*** | 11.0(7.8);n=61 | 12.9(9.5);n=75 | <0.0001*** |
| Hospital stay, days | 26.6(35.0);n=202 | 40.6(36.0);n=244 | <0.0001*** | 33.7(38.8);n=61 | 39.9(39.6);n=75 | <0.0001*** |
| *Acute myocardial infarction (N=38)* |  |  |  | *Acute myocardial infarction (N=8)* |  |  |
| Average readmission interval between episodes, days | 498.3(1094.7);n=32 | 42.1(53.5);n=38 | <0.0001*** | 1576.1(2728.8);n=6 | 38.9(16.4);n=8 | <0.0001*** |
| No. of episodes | 6.4(5.7);n=32 | 13.2(8.9);n=38 | <0.0001*** | 5.8(3.9);n=6 | 17.4(11.6);n=8 | <0.0001*** |
| Hospital stay, days | 24.5(28.9);n=32 | 34.3(30.4);n=38 | <0.0001*** | 11.5(6.4);n=6 | 20.8(12.3);n=8 | <0.0001*** |
| *Atrial fibrillation (N=54)* |  |  |  | *Atrial fibrillation (N=8)* |  |  |
| Average readmission interval between episodes, days | 383.9(905.9);n=46 | 43.4(48.0);n=54 | <0.0001*** | 715.5(1861.5);n=6 | 49.4(45.0);n=8 | <0.0001*** |
| No. of episodes | 9.2(8.8);n=46 | 12.5(10.2);n=54 | <0.0001*** | 15.6(11.1);n=6 | 9.0(5.0);n=8 | <0.0001*** |
| Hospital stay, days | 28.7(29.3);n=46 | 43.6(54.6);n=54 | <0.0001*** | 30.4(27.5);n=6 | 23.4(25.5);n=8 | <0.0001*** |
| *Atrial flutter (N=6)* |  |  |  | *Atrial flutter (N=2)* |  |  |
| Average readmission interval between episodes, days | 190.2(175.8);n=5 | 42.5(47.1);n=6 | <0.0001*** | - | - | - |
| No. of episodes | 3.8(2.2);n=5 | 11.5(7.8);n=6 | <0.0001*** | - | - | - |
| Hospital stay, days | 28.0(32.5);n=5 | 33.3(22.1);n=6 | <0.0001*** | - | - | - |

IR: Incidence rate;
PD-1: Programmed death-1 inhibitors;
PD-L1: programmed death-ligand 1 inhibitors.

### Sensitivity analysis

**Supplementary Table 7** presented the adjusted hazard ratios (and 95% CIs) of PD-L1 v.s. PD-1 with cause-specific and subdistribution hazard competing risk analysis models for new onset cardiac composite and mortality outcomes after 1:2 propensity score matching. **Supplementary Table 8** presented the hazard ratios for associations of PD-L1 v.s. PD-1 using Cox proportional hazard model for adverse new onset cardiac composite and mortality outcome in the 1:2 matched cohort, with half-year lag time. **Supplementary Table 9** presented the risk of incident new onset cardiac composite and mortality outcomes associated with treatment of PD-L1 v.s. PD-1 with multiple matching adjustment approaches including propensity score stratification, high-dimensional propensity score matching, and propensity score matching with inverse probability of treatment weighting. The above analysis confirmed the protective effects of PD-L1 treatment over PD-1 treatment on new onset cardiac complications and mortality risks (HR<1, P value<0.05).

### Prediction strength of subclinical inflammatory biomarkers

In the matched cohort, higher aspartate transaminase-to-alanine transaminase ratio (HR: 1.09, 95% CI: [1.04-1.14], P value=0.0004) and higher monocyte-to-lymphocyte ratio (HR: 1.2, 95% CI: [1.02-1.43], P value=0.0305) were significantly associated with all-cause mortality. A higher urea-to-creatinine ratio (HR: 1.01, 95% CI: [1.00-1.01], P value=0.0007) was significantly associated with new onset cardiac composite outcome (Table 4). The boxplots of inflammatory biomarkers stratified by PD-1/PD-L1 inhibitor use and development of adverse outcomes are shown in Figure 4.

**Figure 4.**
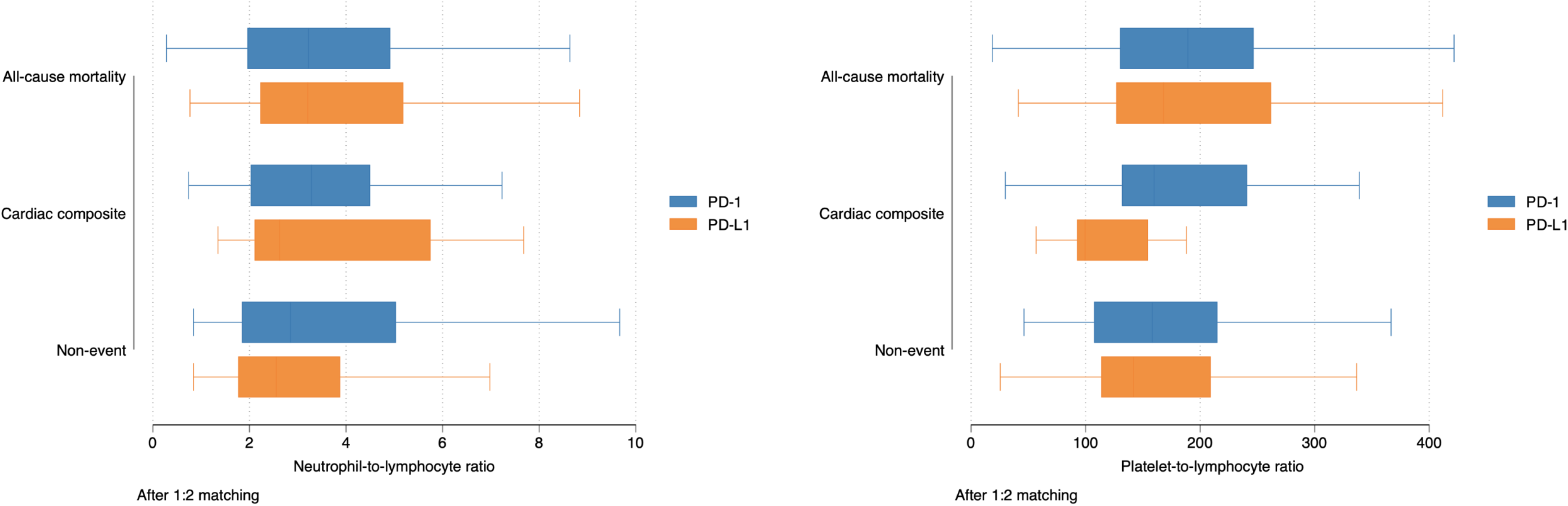
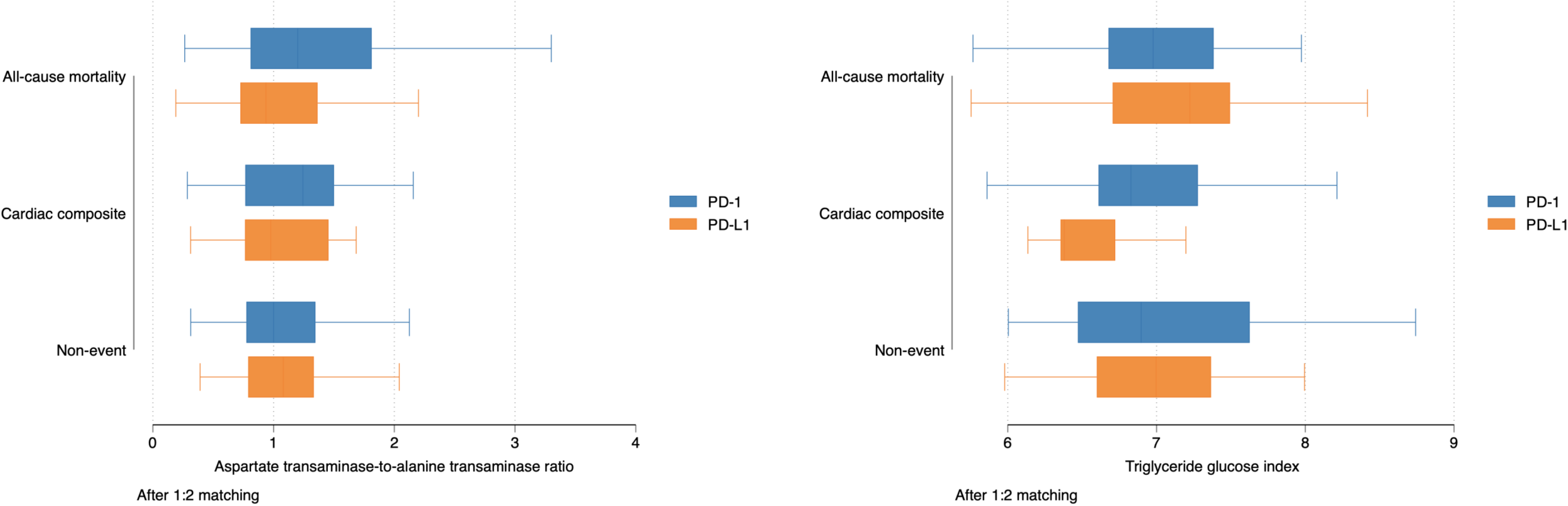
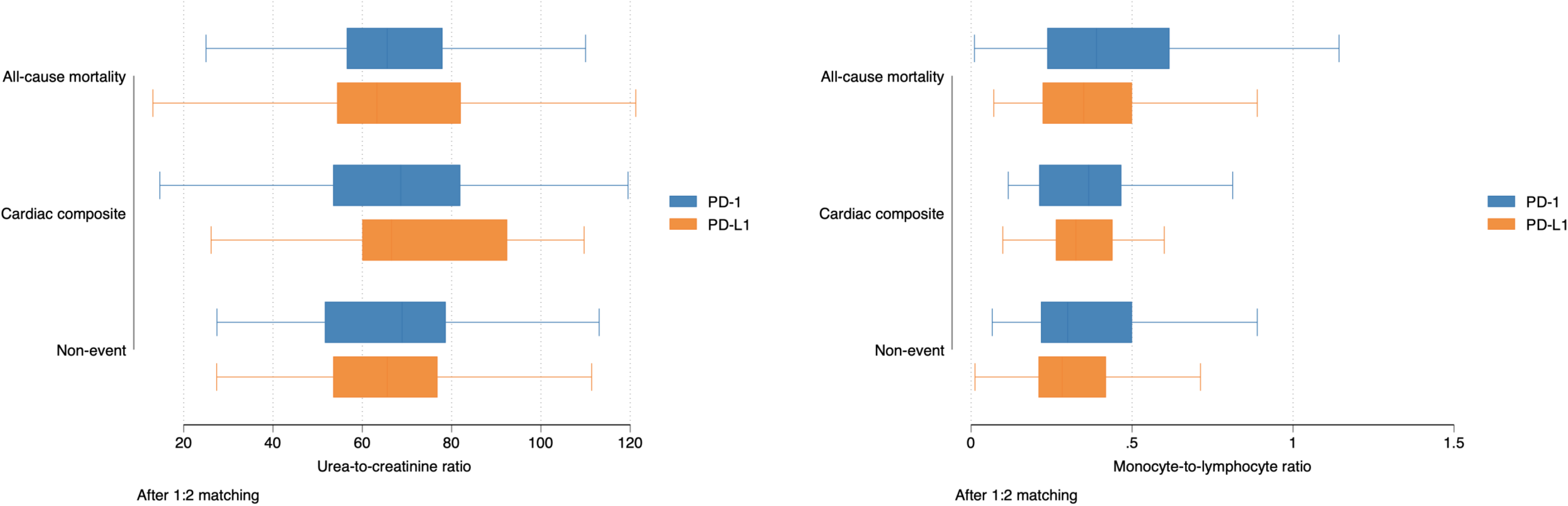
Boxplots of subclinical inflammatory biomarkers stratified by PD-1/PD-L1 inhibitor use and development of the composite outcome in the matched cohort.

**Figure 5.**
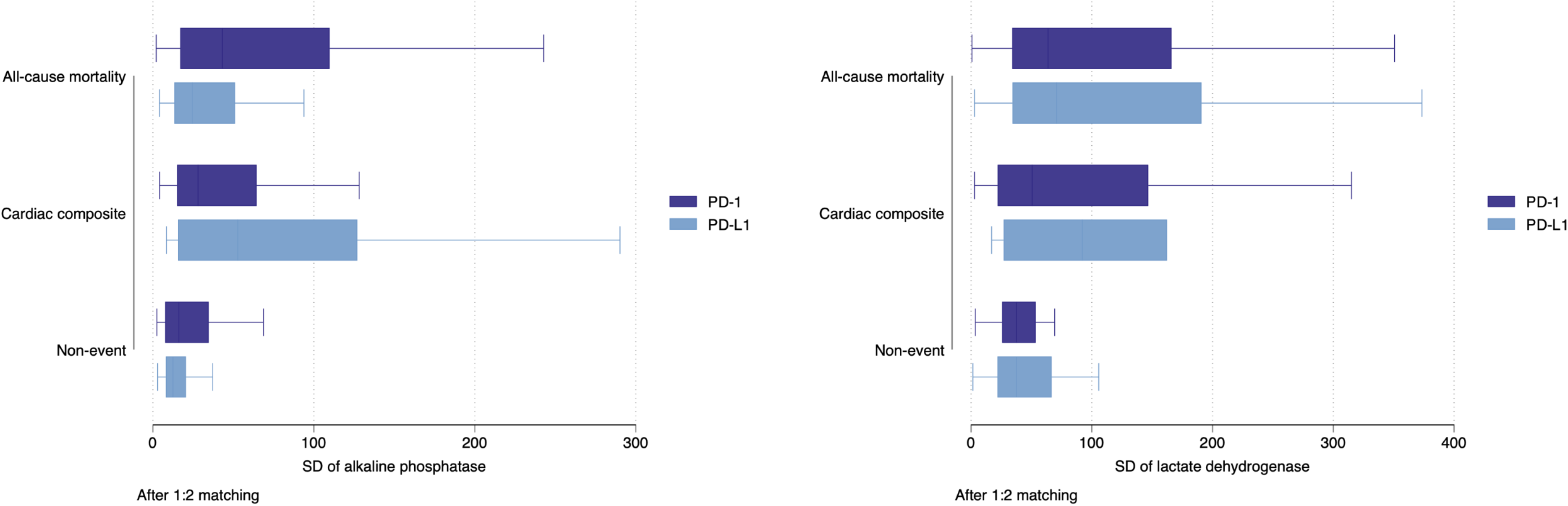
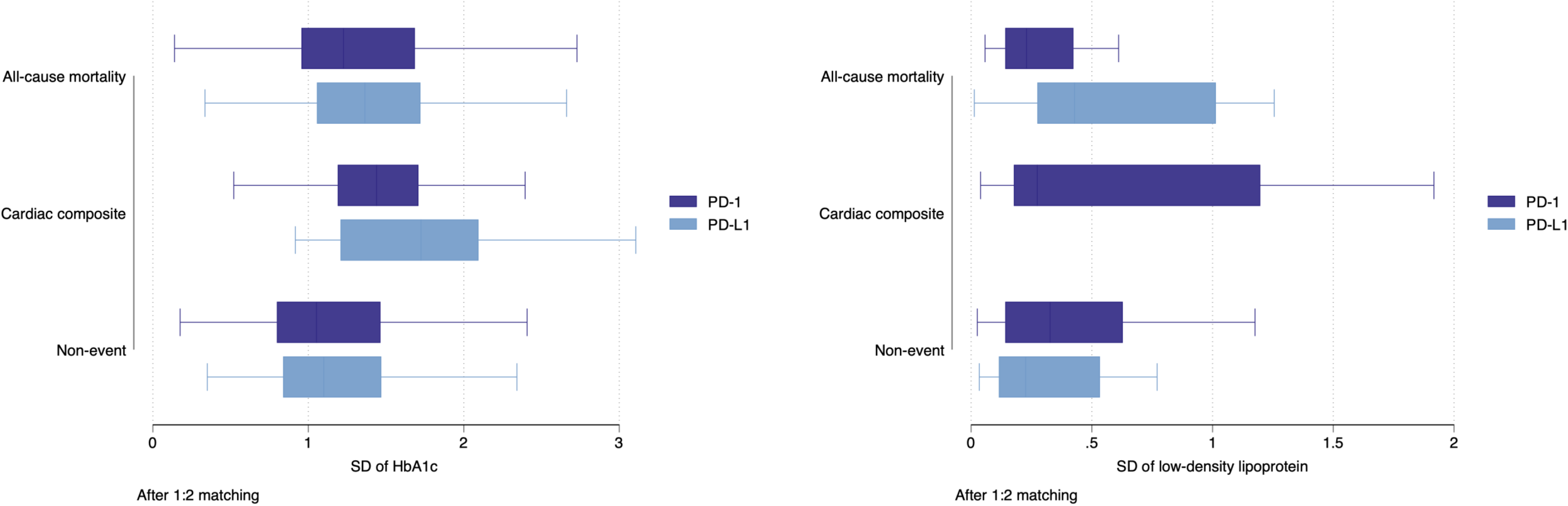
Boxplots of significant laboratory examination variability stratified by PD-1/PD-L1 inhibitor use and development of the composite outcome in the matched cohort.

## Discussion

The main findings are that: i) the incidence of cardiovascular complications after PD-1 or PD-L1 inhibitor use was 16% in this territory-wide cohort of Chinese patients from Hong Kong, ii) multivariate Cox regression showed older age, a shorter average readmission interval and a higher number of hospital admissions were significant predictors of cardiovascular complications and iii) patients who developed cardiovascular complications had shorter average readmission interval and higher number of hospitalizations after treatment with PD-1/PD-L1 inhibitors.

Cardiac involvement in PD1 or PD-L1 inhibitors is variable, and can potentially affect the conduction system, myocardium or pericardium (16). Thus, heart block (17), Takotsubo cardiomyopathy (18), myocarditis (19, 20) and pericarditis (21) have been reported. A meta-analysis performed in 2018 found that anti-PD-1/PD-L1-related fatalities were often from pneumonitis (333 [35%]), hepatitis (115 [22%]), and neurotoxic effects (50 [15%]). Combination PD-1/ cytotoxic T-lymphocyte-associated protein 4 (CTLA-4) deaths were frequently from colitis (32 [37%]) and myocarditis (22 [25%]) (22). In an analysis of the World Health Organization global database of adverse drug reactions in 2019, 2.1% of 106025 patients receiving PD-1 or PD-L1 inhibitors had cardiovascular complications (8). However, previous studies have largely been limited to case reports (17, 23), case series (24), single-center studies (25) or small registries (26, 27). In this territory-wide study from Hong Kong, we found that cardiovascular complications occurred in 16% of all patients receiving PD1 or PD-L1 inhibitors. Of these, the commonest is heart failure. Previously, acute heart failure has been described in the context of myocarditis (28), but heart failure without myocarditis has also been reported (29). Our study also identified cases of acute myocardial infarction following the initiation of PD-1/PD-L1 inhibitor therapy. Such findings would be in keeping with coronary toxicity that has been reported in the context of PD-1 inhibitor therapy (26).

Interestingly, our study did not identify any patients with myocarditis after treatment with PD-1 or PD-L1 inhibitors. Moreover, within the excluded patients with prior cardiovascular complications, none developed subsequent myocarditis. In 964 patients attending Massachusetts General Hospital, the incidence of myocarditis was 1.1% (n = 35) (27). In this cohort, myocarditis was more frequently observed in patients with pre-existing cardiovascular comorbidities. Nevertheless, another study using the VigiBase database found 101 cases of severe myocarditis, of which 75% of the myocarditis cases did not have pre-existing cardiovascular disease (30). A single-center study of 283 patients from China found only 3 cases (1.1%) of myocarditis, with variable presentations such as palpitations, dyspnea, and fatigue, or asymptomatic with incidental finding of grade 3 atrioventricular block and premature ventricular complexes on the electrocardiogram (25). In a pooled, retrospective review of three trials including 448 patients with advanced melanoma receiving PD-1/PD-L1 inhibitor therapy, no cases of myocarditis were identified (31). In association with myocarditis, different investigators have reported the presence of conduction abnormalities in the form of atrioventricular block (17,24,25,32).

## Limitations

There are some limitations of this study that should be acknowledged. Firstly, this was an administrative database study, and therefore cancer staging details could not be extracted. Secondly, under-coding or miscoding remains a possibility as with studies of a similar nature.

## Conclusions

Compared with PD-1 inhibitor use, PD-L1 inhibitor use was significantly associated with lower risks of cardiac complications and all-cause mortality both before and after propensity score matching. Patients who developed cardiovascular complications had shorter average readmission intervals and a higher number of hospitalizations after treatment with PD-1/PD-L1 inhibitors.

## Data Availability

Anonymized dataset available upon request for data checking, further research or validation purposes.

## Consent for publication

N/A.

## Availability of data and material

Data availability upon request to the corresponding author.

## Authors’ contributions

JZ, SL, GT: data analysis, data interpretation, statistical analysis, manuscript drafting, critical revision of manuscript

IL, SL, LY, TL, YX, YZ: data interpretation, manuscript drafting

EWC, ICKW: project planning, data acquisition, data interpretation, critical revision of manuscript

QZ: study conception, study supervision, project planning, data interpretation, statistical analysis, manuscript drafting, critical revision of manuscript

## Acknowledgements

None.

