## Supplementary Appendix for "Adverse Cardiovascular Complications Following Prescription of Programmed Cell Death 1 (PD-1) and Programmed Cell Death Ligand 1 (PD-L1) Inhibitors: A Propensity-Score Matched Cohort Study with Competing Risk Analysis"

**Supplementary Table 1. PD-1 and PD-L1 inhibitor drugs.**

| **PD-1 inhibitors** | **PD-L1 inhibitors** |
|---|---|
| Pembrolizumab | Atezolizumab (Tecentriq) |
| Nivolumab | Avelumab (Bavencio) |
| Cemiplimab | Durvalumab (Imfinzi) |
| Spartalizumab | KN035 |
| Camrelizumab | CK-301 |
| Sintilimab (IBI308) | AUNP12 |
| Tislelizumab (BGB-A317) | CA-170 |
| Toripalimab (JS 001) | BMS-986189 |
| Dostarlimab (TSR-042, WBP-285) | |
| INCMGA00012 (MGA012) | |
| AMP-224 | |
| AMP-514 (MEDI0680) | |

**Supplementary Table 2. Codes for comorbidities.**

| | |
|---|---|
| Diabetes mellitus | 250 250.01 250.02 250.03 250.1 250.11 250.12 250.13 250.2 250.21 250.22 250.23 250.3 250.31 250.32 250.33 250.4 250.41 250.42 250.43 250.5 250.51 250.52 250.53 250.6 250.61 250.62 250.63 250.7 250.71 250.72 250.73 250.8 250.81 250.82 250.83 250.9 250.91 250.92 250.93 |
| Hypertension | 401 401.1 401.9 402 402.01 402.1 402.11 402.9 402.91 403 403.01 403.1 403.11 403.9 403.91 404 404.01 404.02 404.03 404.1 404.11 404.12 404.13 404.9 404.91 404.92 404.93 405 405.01 405.09 405.1 405.11 405.19 405.9 405.91 405.99 437.2 |
| Heart failure | 428 428 428.1 428.2 428.2 428.21 428.22 428.23 428.3 428.3 428.31 428.32 428.33 428.4 428.4 428.41 428.42 428.43 |

| | |
|---|---|
| | 428.9 398.91 402.01 402.11 402.91 404.01 404.03 404.11 404.13 404.91 404.93 |
| Atrial fibrillation | 427.31 429.4 |
| Maligt dysrhythmia | 426 426.12 426.13 426.51 426.52 426.54 427.1 427.4 427.41 427.42 427.5 |
| Atrial flutter | 427.32 |
| Liver diseases | 456 456.1 456.2 572.2 572.3 572.4 572.8 571.4 571.5 571.6 |
| Chronic obstructive pulmonary disease | 490 491 492 493 494 495 496 491.1 491.2 491.21 491.22 491.8 491.9 492.8 493.01 493.02 493.1 493.11 493.12 493.2 493.21 493.22 493.8 493.81 493.82 493.9 493.91 493.92 494.1 495.1 495.2 495.3 495.4 495.5 495.6 495.7 495.8 495.9 |
| Gastrointestinal bleeding | 531 531.2 531.4 531.6 532 532.2 532.4 532.6 533 533.2 533.4 533.6 534 534.2 534.4 534.6 535.01 535.11 535.21 535.31 535.41 535.51 535.61 535.71 562.02 562.03 562.12 562.13 569.3 569.85 569.86 578 578.1 578.9 |
| Hip fractures/accident falls | 805 805 805 805.01 805.02 805.03 805.04 805.05 805.06 805.07 805.08 805.1 805.1 805.11 805.12 805.13 805.14 805.15 805.16 805.17 805.18 805.2 805.3 805.4 805.5 805.6 805.7 805.8 805.9 812 812 812 812.01 812.02 812.03 812.09 812.1 812.1 812.11 812.12 812.13 812.19 812.2 812.2 812.21 812.3 812.3 812.31 812.4 812.4 812.41 812.42 812.43 812.44 812.49 812.5 812.5 812.51 812.52 812.53 812.54 812.59 813 813 813 813.01 813.02 813.03 813.04 813.05 813.06 813.07 813.08 813.1 813.1 813.11 813.12 813.13 813.14 813.15 813.16 813.17 813.18 813.2 813.2 813.21 813.22 813.23 813.3 813.3 813.31 813.32 813.33 813.4 813.4 813.41 813.42 813.43 813.44 813.45 813.46 813.47 813.5 813.5 813.51 813.52 813.53 813.54 813.8 813.8 813.81 813.82 813.83 813.9 813.9 813.91 813.92 813.93 814 814 814 814.01 814.02 814.03 814.04 814.05 814.06 814.07 814.08 814.09 814.1 814.1 814.11 814.12 814.13 814.14 814.15 814.16 814.17 814.18 814.19 820 820 820 820.01 820.02 820.03 820.09 820.1 820.1 820.11 820.12 820.13 820.19 820.2 820.2 820.21 820.22 820.3 820.3 820.31 820.32 820.8 820.9 170 170 170.1 170.2 170.3 170.4 170.5 170.6 170.7 170.8 170.9 710 710.1 710.4 714 714.1 714.2 714.81 725 E880 |

| |
|---|
| E880.0 E880.1 E880.9 E881 E881.0 E881.1 E882 E883 E883.0 E883.1 E883.2 E883.9 E884 E884.0 E884.1 E884.2 E884.3 E884.4 E884.5 E884.6 E884.9 E885 E885.0 E885.1 E885.2 E885.3 E885.4 E885.9 E886 E886.0 E886.9 E887 E888 E888.0 E888.1 E888.8 E888.9 |
| Malignant dysrhythmia 426 426.12 426.13 426.51 426.52 426.54 427.1 427.4 427.41 427.42 427.5 |
| Renal diseases 582 582 582.1 582.2 582.4 582.8 582.81 582.89 582.9 583 583 583.1 583.2 583.4 583.6 583.7 585 585.1 585.2 585.3 585.4 585.5 585.6 585.9 586 588 588 588.1 588.8 588.81 588.89 588.9 |
| Endocrine diseases 259 259 259.1 259.2 259.3 259.4 259.5 259.5 259.51 259.52 259.8 259.9 |
| Peripheral vascular disease 250.7 443.9 443 443.1 443.2 443.21 443.22 443.23 443.24 443.29 443.8 443.81 443.82 443.89 441 443.9 785.4 V43.4 |
| Stroke/transient ischemic attack 435 435.1 435.2 435.3 435.8 435.9 433.81 433.91 434 436 437 437.1 433.31 433.01 434.01 434.1 434.11 434.9 434.91 437.2 437.3 437.4 437.5 437.6 437.7 437.8 437.9 |
| Gastrointestinal bleeding 531 531.2 531.4 531.6 532 532.2 532.4 532.6 533 533.2 533.4 533.6 534 534.2 534.4 534.6 535.01 535.11 535.21 535.31 535.41 535.51 535.61 535.71 562.02 562.03 562.12 562.13 569.3 569.85 569.86 578 578.1 578.9 |
| Ischemic heart disease 410.01 410.02 410.1 410.11 410.12 410.2 410.21 410.22 410.3 410.31 410.32 410.4 410.41 410.42 410.5 410.51 410.52 410.6 410.61 410.62 410.7 410.71 410.72 410.8 410.81 410.82 410.9 410.91 410.92 411 411.1 411.8 411.81 411.89 413 413.1 413.9 414 414.01 414.02 414.03 414.04 414.05 414.06 414.07 414.1 414.11 414.12 414.19 414.2 414.3 414.4 414.8 414.9 410 412 |
| Cancer 140-239 |

**Supplementary Table 3. Logistic regression analysis of confounding characteristics before conducting propensity score matching (Mortality as outcome).**

| Variables | Coef. | St.Err. | t-value | p-value | [95% Conf | Interval] | Sig |
|---|---|---|---|---|---|---|---|
| PD-L1 v.s. PD-1 | .45 | .067 | -5.35 | 0 | .336 | .603 | *** |
| Male gender | .887 | .096 | -1.11 | .269 | .716 | 1.097 | |
| Baseline age, years | .998 | .004 | -0.46 | .645 | .99 | 1.006 | |
| Charlson standard comorbidity index | 1.11 | .02 | 5.82 | 0 | 1.072 | 1.15 | *** |
| Hypertension | 1.1 | .188 | 0.56 | .577 | .787 | 1.537 | |
| Liver diseases | 1.404 | .253 | 1.88 | .06 | .986 | 1.998 | * |
| Hip fractures/accident falls | 1.335 | .369 | 1.04 | .297 | .776 | 2.295 | |
| Renal diseases | .887 | .127 | -0.84 | .401 | .671 | 1.173 | |
| Diabetes mellitus | .91 | .19 | -0.45 | .651 | .604 | 1.37 | |
| Maligt dysrhythmia | 1.832 | 1.439 | 0.77 | .441 | .393 | 8.545 | |
| Chronic obstructive pulmonary disease | 1.046 | .628 | 0.08 | .94 | .323 | 3.391 | |
| Ischemic heart disease | .771 | .277 | -0.72 | .47 | .381 | 1.56 | |
| Peripheral vascular disease | 2.07 | 1.64 | 0.92 | .359 | .438 | 9.779 | |
| Endocrine diseases | .895 | .107 | -0.92 | .356 | .708 | 1.132 | |
| Gastrointestinal diseases | 1.092 | .121 | 0.79 | .428 | .879 | 1.356 | |
| Stroke/transient ischemic attack | 1.198 | .39 | 0.55 | .58 | .633 | 2.267 | |
| Anticoagulants | .832 | .084 | -1.81 | .07 | .682 | 1.015 | * |

| | | | | | | | |
|---|---|---|---|---|---|---|---|
| Steroids | 1 | . | . | . | . | . | |
| Constant | 1.549 | .388 | 1.75 | .081 | .948 | 2.532 | * |

| | | | |
|---|---|---|---|
| Mean dependent var | 0.673 | SD dependent var | 0.469 |
| Pseudo r-squared | 0.033 | Number of obs | 1959.000 |
| Chi-square | 82.559 | Prob > chi2 | 0.000 |
| Akaike crit. (AIC) | 2428.892 | Bayesian crit. (BIC) | 2529.335 |

*** p<.01, ** p<.05, * p<.1

**Supplementary Table 4. Confounding balancing comparisons of PD-L1 users and PD-1 users after propensity score matching with 1:2 nearest neighbor search using Stata.**

| | Mean | | | t-test | | |
|---|---|---|---|---|---|---|
| **Variable** | **Treated** | **Control** | **%bias** | **t** | **p>t** | **V(T)/V(C)** |
| Male gender | 0.747 | 0.729 | 4 | 0.43 | 0.666 | . |
| Baseline age, years | 63.057 | 63.851 | -6.5 | -0.7 | 0.487 | 0.57* |
| Charlson standard comorbidity index | 6.534 | 6.525 | 0.3 | 0.03 | 0.976 | 0.92 |
| Hypertension | 0.131 | 0.12 | 3.4 | 0.36 | 0.72 | . |

| | | | | | | |
|---|---|---|---|---|---|---|
| Liver diseases | 0.036 | 0.041 | -1.8 | -0.25 | 0.805 | . |
| Hip fractures/accident falls | 0.059 | 0.041 | 8.5 | 0.87 | 0.383 | . |
| Renal diseases | 0.131 | 0.143 | -3.2 | -0.35 | 0.73 | . |
| Diabetes mellitus | 0.09 | 0.104 | -4.9 | -0.48 | 0.631 | . |
| Maligt dysrhythmia | 0 | 0 | . | . | . | . |
| Chronic obstructive pulmonary disease | 0.009 | 0.011 | -2.5 | -0.24 | 0.813 | . |
| Ischemic heart disease | 0.023 | 0.018 | 2.8 | 0.34 | 0.737 | . |
| Peripheral vascular disease | 0.005 | 0.005 | 0 | 0 | 1 | . |
| Endocrine diseases | 0.253 | 0.233 | 4.6 | 0.5 | 0.619 | . |
| Gastrointestinal diseases | 0.833 | 0.839 | -1.6 | -0.19 | 0.848 | . |
| Stroke/transient ischemic attack | 0.036 | 0.038 | -1.2 | -0.13 | 0.9 | . |
| Anticoagulants | 0.543 | 0.55 | -1.4 | -0.14 | 0.886 | . |
| Steroids | 0.543 | 0.55 | -1.4 | -0.14 | 0.886 | . |
| * if variance ratio outside [0.77; 1.30] | | | | | | |
| Ps R2 | LR chi2 | p>chi2 | MeanBias | | | |
| 0.005 | 3.03 | 1 | 3 | | | |
| MedBias | B | R | %Var | | | |
| 2.6 | 16.6 | 0.76 | 50 | | | |
| * if B>25%, R outside [0.5; 2] | | | | | | |

**Supplementary Table 5. Estimations of bootstrapped standard error (replications=50) that incorporates the propensity matching with 1:2 nearest neighbor search strategy.**

| **Observed Coef.** | **Bootstrap Std. Err.** | **z** | **P>z** | **Normal based [95% Conf. Interval]** |
|---|---|---|---|---|
| -0.1447964 | 0.0571656 | -2.53 | 0.011 | [-0.2568388, -0.032754] |

**Supplementary Table 6. Clinical characteristics of patients with/without mortality risk before and after 1:2 propensity score matching.**

* for *SMD* 0.2; APTT: applied partial thromboplastin test; PD-1: Programmed death 1 inhibitors; PD-L1: programmed death 1 ligand inhibitors

| **Characteristics** | **Before matching** | | **SMD** | **After 1:2 matching** | | **SMD** |
|---|---|---|---|---|---|---|
| | **All-cause mortality (N=1319)** Mean(SD);N or Count(%) | **Alive (N=640)** Mean(SD);N or Count(%) | | **All-cause mortality (N=425)** Mean(SD);N or Count(%) | **Alive (N=238)** Mean(SD);N or Count(%) | |
| ***Demographics*** | | | | | | |
| Male gender | 892(67.62%) | 449(70.15%) | 0.05 | 318(74.82%) | 180(75.63%) | 0.02 |
| Female gender | 427(32.37%) | 191(29.84%) | 0.05 | 107(25.17%) | 58(24.36%) | 0.02 |
| Baseline age, years | 61.4(13.3);n=1319 | 60.2(14.5);n=640 | 0.09 | 63.2(10.2);n=425 | 62.7(10.2);n=238 | 0.05 |
| <40 | 93(7.05%) | 58(9.06%) | 0.07 | 14(3.29%) | 6(2.52%) | 0.05 |
| [40, 50) | 134(10.15%) | 60(9.37%) | 0.03 | 29(6.82%) | 14(5.88%) | 0.04 |
| [50-60) | 326(24.71%) | 155(24.21%) | 0.01 | 93(21.88%) | 62(26.05%) | 0.1 |
| [60-70) | 421(31.91%) | 210(32.81%) | 0.02 | 186(43.76%) | 102(42.85%) | 0.02 |

| | | | | | | |
|---|---|---|---|---|---|---|
| [70-80) | 265(20.09%) | 126(19.68%) | 0.01 | 87(20.47%) | 46(19.32%) | 0.03 |
| >=80 | 80(6.06%) | 31(4.84%) | 0.05 | 16(3.76%) | 8(3.36%) | 0.02 |
| ***Past comorbidities*** | | | | | | |
| Charlson standard comorbidity index | 6.4(3.3);n=1319 | 5.4(3.1);n=640 | 0.3* | 7.0(3.1);n=425 | 5.6(2.9);n=238 | 0.45* |
| Hypertension | 181(13.72%) | 75(11.71%) | 0.06 | 61(14.35%) | 27(11.34%) | 0.09 |
| Liver diseases | 144(10.91%) | 49(7.65%) | 0.11 | 18(4.23%) | 6(2.52%) | 0.09 |
| Hip fractures/accident falls | 57(4.32%) | 20(3.12%) | 0.06 | 19(4.47%) | 18(7.56%) | 0.13 |
| Renal diseases | 197(14.93%) | 95(14.84%) | <0.01 | 54(12.70%) | 31(13.02%) | 0.01 |
| Diabetes mellitus | 107(8.11%) | 49(7.65%) | 0.02 | 38(8.94%) | 22(9.24%) | 0.01 |
| Maligt dysrhythmia | 10(0.75%) | 2(0.31%) | 0.06 | 0(0.00%) | 2(0.84%) | 0.13 |
| Chronic obstructive pulmonary disease | 11(0.83%) | 4(0.62%) | 0.02 | 5(1.17%) | 1(0.42%) | 0.09 |
| Ischemic heart disease | 40(3.03%) | 21(3.28%) | 0.01 | 8(1.88%) | 7(2.94%) | 0.07 |
| Peripheral vascular disease | 9(0.68%) | 2(0.31%) | 0.05 | 2(0.47%) | 1(0.42%) | 0.01 |
| Endocrine diseases | 377(28.58%) | 171(26.71%) | 0.04 | 111(26.11%) | 55(23.10%) | 0.07 |
| Gastrointestinal diseases | 960(72.78%) | 452(70.62%) | 0.05 | 362(85.17%) | 189(79.41%) | 0.15 |
| Stroke/transient ischemic attack | 56(4.24%) | 24(3.75%) | 0.03 | 17(4.00%) | 7(2.94%) | 0.06 |
| ***Hospitalization*** | | | | | | |

| | | | | | | |
|---|---|---|---|---|---|---|
| Average readmission | 70.8(200.0);n=1247 | 64.0(184.2);n=629 | 0.04 | 72.6(237.2);n=411 | 52.7(118.7);n=236 | 0.11 |
| Total episode number | 12.2(11.8);n=1247 | 18.3(16.6);n=629 | 0.43* | 12.6(10.1);n=411 | 15.9(11.8);n=236 | 0.3* |
| Overall hospital stay, days | 39.3(42.2);n=1247 | 27.1(26.9);n=629 | 0.34* | 38.6(35.2);n=411 | 23.2(19.6);n=236 | 0.54* |
| ***Medications*** | | | | | | |
| PD-L1 v.s. PD-1 | 115(8.71%) | 106(16.56%) | 0.24* | 115(27.05%) | 106(44.53%) | 0.37* |
| PD-L1 expenditure, HKD | 64470.8(52756.4);n=115 | 134899.3(121584.5);n=106 | 0.75* | 64470.8(52756.4);n=115 | 134899.3(121584.5);n=106 | 0.75* |
| Total PD-L1 dose amount, mg | 11141.0(23955.0);n=115 | 14274.9(30903.5);n=106 | 0.11 | 11141.0(23955.0);n=115 | 14274.9(30903.5);n=106 | 0.11 |
| PD-L1 inhibitors duration, days | 100.9(121.4);n=115 | 257.4(236.0);n=106 | 0.83* | 100.9(121.4);n=115 | 257.4(236.0);n=106 | 0.83* |
| PD-1 expenditure | 132406.3(227146.6);n=1204 | 329433.6(364585.4);n=546 | 0.65* | 155528.6(204178.9);n=310 | 307662.2(326498.3);n=144 | 0.56* |
| Total PD-1 dose amount (MG) | 2324.5(9779.2);n=1204 | 3905.8(11851.9);n=546 | 0.15 | 2114.9(5352.1);n=310 | 3186.2(4625.7);n=144 | 0.21* |
| PD-1 inhibitors duration, days | 152.0(200.9);n=1204 | 314.5(272.0);n=546 | 0.68* | 162.9(209.0);n=310 | 304.0(257.8);n=144 | 0.6* |
| Anticoagulants | 727(55.11%) | 381(59.53%) | 0.09 | 233(54.82%) | 129(54.20%) | 0.01 |
| Steroids | 727(55.11%) | 381(59.53%) | 0.09 | 233(54.82%) | 129(54.20%) | 0.01 |
| ***Inflamatory subclinical biomarkers*** | | | | | | |
| Neutrophil-to-lymphocyte ratio | 4.8(6.8);n=1316 | 4.2(5.5);n=636 | 0.09 | 4.6(7.5);n=425 | 3.7(4.0);n=238 | 0.15 |
| Platelet-to-lymphocyte ratio | 220.6(276.2);n=1317 | 195.2(156.2);n=636 | 0.11 | 226.7(365.3);n=425 | 180.8(136.5);n=238 | 0.17 |

| | | | | | | |
|---|---|---|---|---|---|---|
| Aspartate transaminase-to-alanine transaminase ratio | 2.0(4.3);n=888 | 1.4(1.2);n=420 | 0.22* | 1.6(2.5);n=254 | 1.2(0.7);n=151 | 0.24* |
| Triglyceride glucose index | 7.05(0.61);n=394 | 7.07(0.66);n=186 | 0.03 | 7.0(0.6);n=134 | 7.1(0.7);n=73 | 0.07 |
| Urea-to-creatinine ratio | 73.4(39.8);n=1303 | 72.6(42.6);n=634 | 0.02 | 70.2(32.5);n=422 | 68.2(22.1);n=237 | 0.07 |
| Monocyte-to-lymphocyte ratio | 0.5(0.4);n=1314 | 0.4(0.6);n=636 | 0.06 | 0.5(0.4);n=424 | 0.4(0.4);n=238 | 0.19 |
| ***Complete blood counts*** | | | | | | |
| Mean corpuscular volume, fL | 88.1(8.3);n=1317 | 87.8(8.0);n=636 | 0.04 | 88.3(7.6);n=425 | 88.1(8.1);n=238 | 0.02 |
| Eosinophil, x10^9/L | 0.17(0.26);n=1316 | 0.21(0.38);n=636 | 0.12 | 0.2(0.29);n=425 | 0.21(0.21);n=238 | 0.04 |
| Lymphocyte, x10^9/L | 1.5(0.9);n=1317 | 1.6(0.8);n=636 | 0.08 | 1.5(0.8);n=425 | 1.6(0.7);n=238 | 0.14 |
| Metamyelocyte, x10^9/L | 0.4(0.7);n=169 | 1.9(10.0);n=56 | 0.22* | 0.3(0.4);n=57 | 0.5(0.7);n=15 | 0.36* |
| Monocyte, x10^9/L | 0.54(0.3);n=1317 | 0.52(0.33);n=636 | 0.06 | 0.6(0.3);n=425 | 0.5(0.3);n=238 | 0.17 |
| Neutrophil, x10^9/L | 5.1(3.3);n=1317 | 4.9(3.3);n=636 | 0.07 | 5.1(3.1);n=425 | 4.8(3.3);n=238 | 0.1 |
| White blood count, x10^9/L | 7.5(6.2);n=1317 | 7.2(3.6);n=636 | 0.07 | 7.4(3.6);n=425 | 7.1(3.7);n=238 | 0.08 |
| Mean cell haemoglobin, pg | 30.7(3.4);n=1317 | 30.6(3.3);n=636 | 0.01 | 30.7(3.2);n=425 | 30.9(3.2);n=238 | 0.07 |
| Myelocyte, x10^9/L | 0.5(0.9);n=249 | 1.5(6.0);n=79 | 0.24* | 0.4(0.8);n=76 | 1.5(2.6);n=21 | 0.58* |
| Platelet, x10^9/L | 249.1(110.5);n=1317 | 245.6(104.4);n=636 | 0.03 | 259.0(103.0);n=425 | 241.3(90.2);n=238 | 0.18 |
| Red blood count, x10^12/L | 4.4(0.7);n=1317 | 4.5(0.7);n=636 | 0.14 | 4.4(0.7);n=425 | 4.6(0.6);n=238 | 0.22* |
| Hematocrit, L/L | 0.38(0.06);n=1272 | 0.39(0.05);n=623 | 0.15 | 0.39(0.05);n=396 | 0.4(0.05);n=230 | 0.22* |

| | | | | | | |
|---|---|---|---|---|---|---|
| ***Renal and liver functions*** | | | | | | |
| K/Potassium, mmol/L | 4.12(0.44);n=1306 | 4.14(0.43);n=635 | 0.06 | 4.12(0.43);n=423 | 4.15(0.42);n=238 | 0.06 |
| Urate, mmol/L | 0.33(0.15);n=419 | 0.32(0.12);n=208 | 0.04 | 0.34(0.14);n=111 | 0.33(0.1);n=75 | 0.05 |
| Albumin, g/L | 38.7(6.2);n=1305 | 40.2(5.1);n=634 | 0.26* | 39.1(5.7);n=423 | 40.3(4.8);n=237 | 0.22* |
| Na/Sodium, mmol/L | 138.8(4.1);n=1306 | 139.8(3.0);n=635 | 0.29* | 138.9(3.7);n=423 | 139.8(2.7);n=238 | 0.28* |
| Urea, mmol/L | 5.5(2.3);n=1303 | 5.6(2.6);n=634 | 0.02 | 5.2(1.8);n=422 | 5.5(1.7);n=237 | 0.14 |
| Protein, g/L | 71.5(13.6);n=1252 | 73.3(11.0);n=601 | 0.15 | 70.7(15.8);n=400 | 73.4(11.4);n=224 | 0.19 |
| Bilirubin, umol/L | 14.0(28.2);n=1306 | 10.8(17.5);n=635 | 0.14 | 11.1(13.2);n=423 | 11.7(26.5);n=238 | 0.03 |
| Creatinine, umol/L | 85.6(70.1);n=1317 | 83.5(42.1);n=635 | 0.04 | 85.2(76.7);n=424 | 84.2(30.8);n=238 | 0.02 |
| SD of creatinine | 41.9(116.3);n=1312 | 22.0(68.8);n=630 | 0.21* | 33.6(78.2);n=422 | 22.8(88.9);n=236 | 0.13 |
| Aspartate transaminase, U/L | 60.5(119.5);n=936 | 41.4(61.1);n=435 | 0.2* | 42.5(64.4);n=283 | 39.0(72.2);n=160 | 0.05 |
| SD of aspartate transaminase | 74.1(223.3);n=855 | 28.1(66.1);n=395 | 0.28* | 41.0(91.7);n=263 | 28.3(68.4);n=146 | 0.16 |
| Alkaline phosphatase, U/L | 114.2(122.3);n=1306 | 97.3(84.6);n=635 | 0.16 | 103.2(104.3);n=423 | 100.0(91.8);n=238 | 0.03 |
| SD of alkaline phosphatase | 81.5(107.3);n=1298 | 30.4(47.3);n=630 | 0.62* | 75.8(103.9);n=419 | 27.0(41.8);n=236 | 0.62* |
| Alanine transaminase, U/L | 37.6(58.7);n=1257 | 34.6(49.1);n=619 | 0.06 | 30.7(27.5);n=394 | 32.7(32.8);n=229 | 0.07 |
| SD of alanine transaminase | 41.5(93.1);n=1244 | 26.0(63.1);n=611 | 0.2 | 30.7(50.8);n=388 | 23.9(53.0);n=224 | 0.13 |
| ***Lipid, iron and calcium profile*** | | | | | | |
| Total iron-binding capacity, L | 40.8(12.8);n=86 | 41.9(9.9);n=17 | 0.1 | 40.6(13.6);n=17 | 47.4(7.8);n=5 | 0.62* |

| | | | | | | |
|---|---|---|---|---|---|---|
| VitaminB12, pmol/L | 425.7(314.0);n=39 | 560.6(372.1);n=10 | 0.39* | 375.2(321.8);n=14 | 522.5(218.3);n=3 | 0.54* |
| Folate, ng/mL | 21.4(10.1);n=56 | 23.2(10.9);n=13 | 0.18 | 22.5(11.8);n=13 | 15.8(3.9);n=6 | 0.76* |
| Ferritin, pmol/L | 2833.9(5276.2);n=66 | 1598.8(2017.0);n=20 | 0.31* | 2547.3(3542.8);n=14 | 880.3(1198.5);n=8 | 0.63* |
| Calcium, mmol/L | 2.31(0.16);n=794 | 2.33(0.13);n=287 | 0.1 | 2.32(0.15);n=235 | 2.33(0.14);n=105 | 0.03 |
| SD of calcium | 0.1(0.07);n=740 | 0.08(0.04);n=247 | 0.32* | 0.09(0.07);n=221 | 0.08(0.04);n=93 | 0.24* |
| Phosphate, mmol/L | 1.05(0.23);n=703 | 1.07(0.19);n=245 | 0.09 | 1.1(0.2);n=203 | 1.0(0.2);n=83 | 0.11 |
| SD of phosphate | 0.2(0.1);n=595 | 0.1(0.1);n=190 | 0.37* | 0.2(0.1);n=175 | 0.1(0.1);n=66 | 0.21* |
| Inorganic, mmol/L | 6.8(8.8);n=202 | 5.0(7.9);n=57 | 0.21* | 7.2(11.4);n=56 | 4.7(5.8);n=23 | 0.28* |
| SD of inorganic | 2.13(3.39);n=86 | 2.11(3.9);n=27 | 0.01 | 1.0(1.6);n=29 | 0.4(0.6);n=12 | 0.43* |
| ***Glycemic and clotting profile*** | | | | | | |
| Triglyceride, mmol/L | 1.4(1.0);n=409 | 1.5(1.2);n=189 | 0.06 | 1.4(1.0);n=140 | 1.5(1.5);n=74 | 0.08 |
| SD of triglyceride | 0.35(0.53);n=202 | 0.36(0.53);n=92 | 0.02 | 0.4(0.5);n=52 | 0.43(0.7);n=38 | 0.06 |
| Glucose, mmol/L | 6.3(2.4);n=1214 | 6.2(1.9);n=559 | 0.05 | 6.2(2.3);n=397 | 6.4(2.0);n=211 | 0.06 |
| SD of glucose | 1.6(1.3);n=1135 | 1.3(1.2);n=503 | 0.18 | 1.6(1.5);n=372 | 1.3(1.1);n=192 | 0.2 |
| HbA1c, g/dL | 12.7(1.9);n=1317 | 13.0(1.9);n=636 | 0.12 | 12.9(1.8);n=425 | 13.3(1.9);n=238 | 0.22* |
| SD of HbA1c | 1.4(0.5);n=1316 | 1.1(0.5);n=631 | 0.51* | 1.4(0.5);n=425 | 1.2(0.5);n=235 | 0.47* |
| High sensitive troponin-I, ng/L | 457.9(12369.6);n=897 | 15.7(71.2);n=244 | 0.05 | 41.6(359.0);n=285 | 15.7(38.6);n=87 | 0.1 |
| SD of high sensitive | 114.8(1195.8);n=612 | 163.5(1792.6);n=143 | 0.03 | 41.8(237.0);n=185 | 12.7(36.3);n=60 | 0.17 |

| | | | | | | |
|---|---|---|---|---|---|---|
| troponin-I | | | | | | |
| APTT, second | 30.9(5.9);n=528 | 30.6(3.9);n=166 | 0.06 | 30.5(4.0);n=160 | 30.3(3.5);n=57 | 0.04 |
| SD of APTT | 2.7(3.3);n=389 | 2.2(2.5);n=121 | 0.17 | 2.0(1.7);n=116 | 2.2(2.0);n=36 | 0.06 |
| Lactate dehydrogenase, U/L | 362.2(492.6);n=1014 | 231.6(95.5);n=417 | 0.37* | 338.9(420.8);n=318 | 223.0(72.8);n=148 | 0.38* |
| SD of lactate dehydrogenase | 195.0(461.6);n=819 | 53.5(48.1);n=321 | 0.43* | 155.5(239.4);n=252 | 45.0(37.5);n=113 | 0.64* |
| Total cholesterol, mmol/L | 4.57(1.07);n=410 | 4.6(1.01);n=189 | 0.02 | 4.7(1.2);n=140 | 4.5(0.9);n=74 | 0.25* |
| SD of total cholesterol | 0.5(0.4);n=202 | 0.4(0.4);n=93 | 0.18 | 0.6(0.5);n=55 | 0.4(0.4);n=38 | 0.28* |
| Low-density lipoprotein, mmol/L | 2.62(0.94);n=399 | 2.62(0.88);n=184 | <0.01 | 2.8(1.1);n=137 | 2.5(0.7);n=71 | 0.32* |
| SD of low-density lipoprotein | 0.43(0.37);n=192 | 0.38(0.37);n=91 | 0.13 | 0.44(0.44);n=51 | 0.37(0.32);n=37 | 0.17 |
| High-density lipoprotein, mmol/L | 1.3(0.4);n=405 | 1.4(0.4);n=188 | 0.06 | 1.31(0.37);n=139 | 1.32(0.39);n=73 | 0.01 |
| SD of high-density lipoprotein | 0.2(0.1);n=190 | 0.1(0.1);n=90 | 0.33* | 0.2(0.2);n=49 | 0.1(0.1);n=38 | 0.19 |

**Supplementary Table 7. Sensitivity analysis 1: Adjusted hazard ratios (and 95% CIs) of PD-L1 v.s. PD-1 with cause-specific and subdistribution hazard competing risk analysis models for new onset cardiac composite and mortality outcomes after 1:2 propensity score matching.**

* for p≤ 0.05, ** for p ≤ 0.01, *** for p ≤ 0.001; HR: hazard ratio; CI: confidence interval.

| **Model** | **Adverse outcomes** | **PD-L1 v.s. PD-1**<br>**HR [95% CI];p value** |
|---|---|---|
| Cause-specific hazard model | New onset cardiac composite | 0.30[0.16, 0.57]; 0.0002*** |
| | All-cause mortality | 0.93[0.74- 1.17];0.525 |
| Subdistribution hazard model | New onset cardiac composite | 0.35[0.19, 0.55]; <0.0001*** |
| | All-cause mortality | 0.85[0.70- 0.93];0.0312* |

**Supplementary Table 8. Sensitivity analysis 2: Hazard ratios for associations of PD-L1 v.s. PD-1 using Cox proportional hazard model for adverse new onset cardiac composite and mortality outcome in the 1:2 matched cohort, with half-year lag time.**

* for p≤ 0.05, ** for p ≤ 0.01, *** for p ≤ 0.001; HR: hazard ratio; CI: confidence interval.

| **Adverse outcomes** | **PD-L1 v.s. PD-1**<br>**HR [95% CI];P value** |
|---|---|
| New onset cardiac composite | 0.33[0.19, 0.59]; <0.0001*** |
| All-cause mortality | 0.83[0.75- 0.97];0.0225* |

**Supplementary Table 9. Sensitivity analysis 3: Risk of incident new onset cardiac composite and mortality outcomes associated with treatment of PD-L1 v.s. PD-1 with multiple matching adjustment approaches.**

* for p≤ 0.05, ** for p ≤ 0.01, *** for p ≤ 0.001; HR: hazard ratio; CI: confidence interval;

PS: propensity score, HDPS: high dimensional propensity score, IPTW: inverse probability of treatment weighting.

| **Outcome** | **HR after PS stratification** <br> **[95% CI];P value** | **HR after HDPS matching** <br> **[95% CI];P value** | **HR after PS IPTW** <br> **[95% CI]; P value** |
|---|---|---|---|
| Composite outcome | 0.34[0.17, 0.67]; 0.0003*** | 0.36[0.13, 0.65]; <0.0001*** | 0.34[0.16, 0.72]; 0.0001*** |
| All-cause mortality | 0.82[0.63- 0.95];0.0325* | 0.81[0.54- 1.00];0.0045** | 0.80[0.44- 0.96];0.0225* |

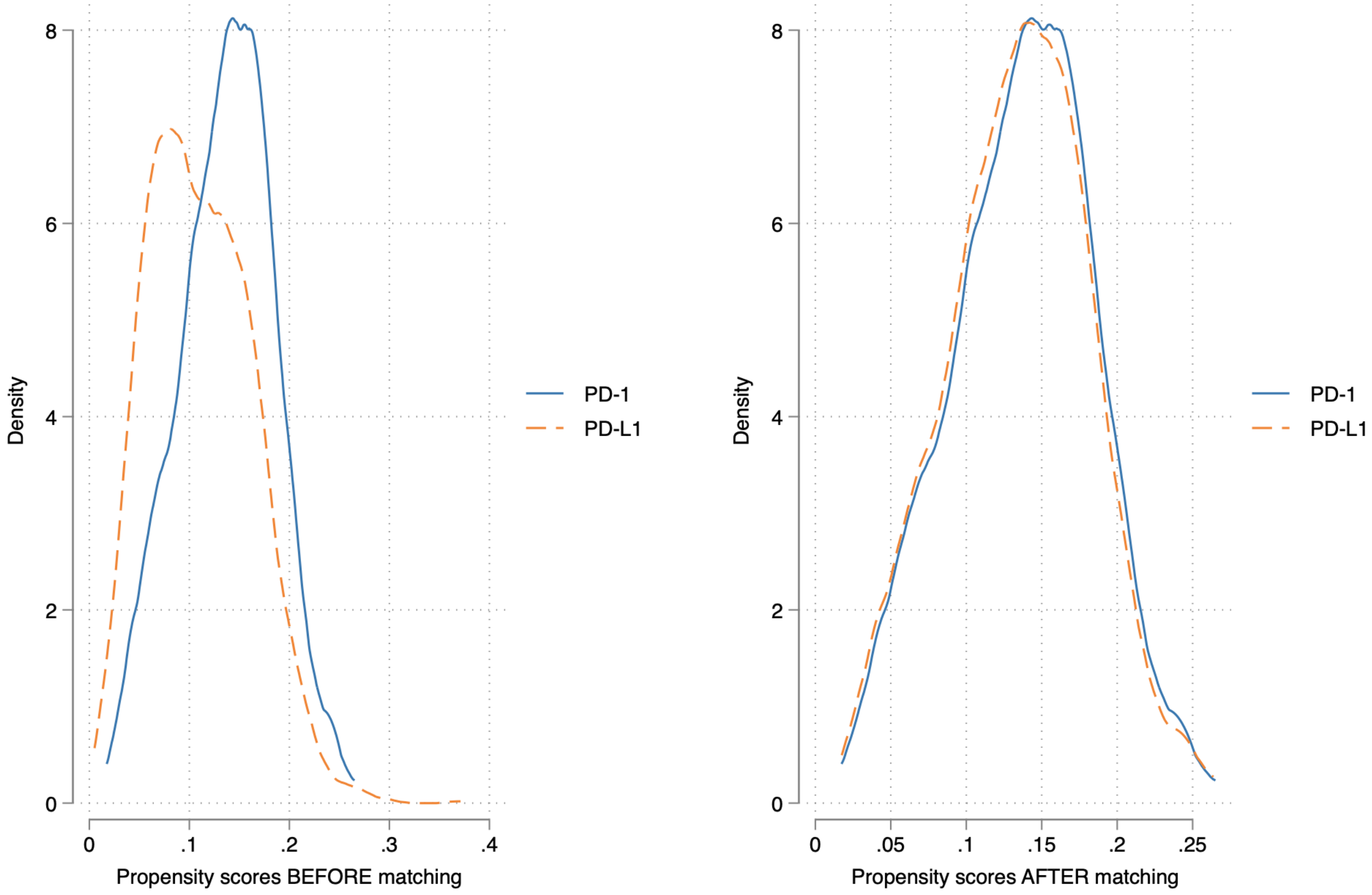


**Supplementary Figure 1. Propensity score matching for PD-L1v.s. PD-1 before and after 1:2 matching with nearest neighbor search strategy.**
